# The cost of primary care consultations associated with long COVID in non-hospitalised adults: a retrospective cohort study using UK primary care data

**DOI:** 10.1101/2023.03.12.23287049

**Authors:** Jake Tufts, Dawit T Zemedikun, Anuradhaa Subramanian, Naijie Guan, Krishna Gokhale, Puja Myles, Tim Williams, Tom Marshall, Melanie Calvert, Karen Matthews, Krishnarajah Nirantharakumar, Louise Jackson, Shamil Haroon

**Affiliations:** University Hospitals of Morecambe Bay NHS Foundation Trust, Lancashire, UK; School of Population and Global Health (M431), The University of Western Australia, 35 Stirling Highway, Perth, WA 6009, Australia; Institute of Applied Health Research, University of Birmingham, Edgbaston, Birmingham, UK; Clinical Practice Research Datalink, Medicines and Healthcare products Regulatory Agency, London; Birmingham Health Partners Centre for Regulatory Science and Innovation, University of Birmingham, Birmingham, UK; National Institute for Health Research (NIHR), Applied Research Collaboration (ARC) West Midlands, Birmingham, UK; NIHR Birmingham Biomedical Research Centre, University Hospital Birmingham and University of Birmingham, Birmingham, UK; NIHR Birmingham-Oxford Blood and Transplant Research Unit (BTRU) in Precision Transplant and Cellular Therapeutics, University of Birmingham, Birmingham, UK; Long Covid SOS, Charity registered in England & Wales, 11A Westland Road, Faringdon, Oxfordshire, UK

## Abstract

**Objectives:** To assess incremental costs of primary care consultations associated with post-Covid-19 condition or long COVID, to estimate associated national costs for the United Kingdom population, and to assess risk factors associated with increased costs.

**Design:** A retrospective cohort study using a propensity score matching approach with an incremental cost method to estimate primary care consultation costs associated with long COVID.

**Setting:** UK based primary care general practitioner (GP), nurse and physiotherapist consultation data from the Clinical Practice Research Datalink Aurum primary care database from 31^st^ January 2020 to 15^th^ April 2021.

**Participants:** 472,173 non-hospitalised adults with confirmed SARS-CoV-2 infection were 1:1 propensity score matched to a pool of eligible patients with the same index date, the same number of prior consultations, and similar background characteristics, but without a record of COVID-19. Patients diagnosed with Long COVID (3,871) and those with World Health Organisation (WHO) defined symptoms of long COVID (30,174) formed two subgroups within the cohort with confirmed SARS-CoV-2 infection.

**Main outcome measures:** Costs were calculated using a bottom-up costing approach with consultation cost per working hour in pound sterling (£) obtained from the Personal Social Services Research Unit’s Unit Costs of Health and Social Care 2021. The average incremental cost in comparison to patients with no record of COVID-19 was produced for each patient group, considering only consultation costs at least 12 weeks from the SARS-CoV-2 infection date or matched date for the comparator group (from 15^th^ April 2020 to 15^th^ April 2021). A sensitivity analysis was undertaken which restricted the study population to only those who had at least 24 weeks of follow-up. National costs were estimated by extrapolating incremental costs to the cumulative incidence of COVID-19 in the UK Office for National Statistics COVID-19 Infection Survey. The impacts of risk factors on the cost of consultations beyond 12 weeks from SARS-CoV-2 infection were assessed using an econometric ordinary least squares (OLS) regression model, where coefficients were interpreted as the percentage change in cost due to a unit increase in the specific factor.

**Results:** The incremental cost of primary care consultations potentially associated with long COVID was £2.44 per patient with COVID-19 per year. This increased to £5.72 in the sensitivity analysis. Extrapolating this to the UK population produced a cost estimate of £23,382,452 (90% credible interval: £21,378,567 to £25,526,052) or £54,814,601 (90% credible interval: £50,116,967 to £59,839,762) in the sensitivity analysis. Among patients with COVID-19 infection, a long COVID diagnosis and longer-term reporting of symptoms were associated with a 43% and 44% increase in primary care consultation costs respectively, compared to patients without long COVID symptoms. Older age (49% relative increase in costs in those aged 80 years or older compared to those aged 18 to 29 years), female sex (4% relative increase in costs compared to males), obesity (4% relative increase in costs compared to those of normal weight), comorbidities and the number of prior consultations were all associated with an increase in the cost of primary care consultations. By contrast, those from black ethnic groups had a 6% reduced relative cost compared to those from white ethnic groups.

**Conclusions:** The costs of primary care consultations associated with long COVID in non-hospitalised adults are substantial. Costs are significantly higher among those diagnosed with long COVID, those with long COVID symptoms, older adults, females, and those with obesity and comorbidities.

**What is already known on this topic?:**

- Long COVID is a global public health challenge, with millions of people affected worldwide.
- People with a history of long COVID use health services, including primary care, at a higher rate than uninfected individuals even beyond the period of acute infection.
- The cost of this increased healthcare use is unknown, impeding planning and forecasting of resource requirements needed to adequately support people with long COVID.

**What this study adds?:**

- Beyond 12 weeks from acute infection, non-hospitalised adults with a history of SARS-CoV-2 infection cost primary care services an additional £2.44 per patient per year greater on average than patients with no prior evidence of infection.
- Due to the high incidence of COVID-19, this represents a substantial cost to primary care services, in the UK exceeding £20 million for consultations associated with long COVID.
- These incremental costs are greater in those with a formal diagnosis of long COVID, those reporting related symptoms, older adults, females, and those with obesity.

## Introduction

Post COVID-19 condition or long COVID is one of the largest public health challenges associated with the COVID-19 pandemic. The World Health Organisation defines it as an illness that occurs following probable or confirmed Severe Acute Respiratory Syndrome Coronavirus-2 (SARS-CoV-2) infection, usually within three months of the acute infection, with symptoms and health effects lasting for at least two months, that cannot be explained by an alternative diagnosis.^1, 2^ The prevalence of long COVID in the UK and worldwide is high.^3^ In June 2022, two million people were estimated to be experiencing self-reported long COVID in the UK alone.^3^ At the time of the current study, over 630 million people worldwide had cumulatively had COVID-19^4^ and 6.2% were estimated to have experienced symptoms lasting beyond three months from infection,^5^ suggesting a global long COVID prevalence of approximately 40 million cases. This burden has steadily increased over the course of the pandemic and of those self-reporting long COVID, 72% reported that their symptoms were adversely affecting their day-to-day activities.^6^

Research has shown that, in comparison to uninfected individuals, those with a history of COVID-19 had significantly higher GP consultation rates post-infection, the vast majority of whom were not hospitalised.^7, 8^ It is therefore likely that long COVID has also led to increased primary care costs but no robust evidence on this has currently been published. Estimating the economic cost of primary care consultations attributed to long COVID can help inform understanding of the economic burden of the condition on health services. Analysing how the costs vary across population subgroups and how they are influenced by risk factors can inform healthcare policy and decisions relating to resource allocation.

The aim of the study was to estimate the excess primary care costs associated with consultations to support non-hospitalised people with long COVID. The three objectives were to estimate the incremental costs of these consultations per patient with a history of COVID-19 beyond 12 weeks from infection, to estimate the national primary care costs of these consultations in the UK, and to assess the association between demographic and clinical risk factors with incremental costs among those with a history of SARS-CoV-2 infection.

This study aimed to estimate the cost of long COVID from a primary care perspective, by quantifying the direct healthcare costs from primary care consultations that can be attributed to supporting people with long COVID, compared to a closely matched cohort of individuals with no record of suspected or confirmed COVID-19.^9^

## Methods

### Study design

A retrospective matched cohort study was conducted using data from a large primary care database based in the UK. The study compared the frequency and costs of primary care consultations in a cohort of individuals with confirmed SARS-CoV-2 infection, at least 12 weeks after infection (representing the longer-term effects of COVID-19 or post COVID-19 condition/long COVID), to a propensity score matched cohort of individuals with no evidence of suspected or confirmed COVID-19. The costs attributed to additional primary consultations to support those with long COVID were estimated for the UK. Healthcare resource use was calculated using a bottom-up approach, and incremental costs were estimated using the matched control method.^10, 11^ The association between patient characteristics and primary care consultation costs among those with confirmed SARS-CoV-2 infection were then assessed. This analysis was part of the Therapies for Long COVID in non-hospitalised individuals (TLC) Study.^12^

### Data source

Data were obtained from the Clinical Practice Research Datalink (CPRD) Aurum database from 31^st^ January 2020 to 15^th^ April 2021.^13^ CPRD Aurum contains anonymised routinely collected data from UK general practices that use the EMIS Web® patient record system software.^14^ In June 2021, over 13 million actively registered patients were included in CPRD Aurum, covering approximately 20% of the UK population and 15% of all general practices in the UK.^13^ The database is representative of the UK population and captures data on patient demographics, diagnoses, symptoms and more. SNOMED CT terms were used for coding diagnoses and symptoms. ^12, 15^ Data extraction was performed using the Data Extraction for Epidemiological Research (DExtER) tool for automated clinical epidemiological studies.^16^

### Study population

Patients were sampled from general practices that were eligible if they had provided research quality data for at least 12 months before the study start date (31^st^ January 2020). Patients were eligible if they were 18 years or older on the study start date, had been registered with a general practice for more than 12 months, and had a minimum of 12 weeks of follow-up. The latter eligibility criterion was included as long COVID is generally defined as symptoms persisting beyond 12 weeks of infection so a minimum of 12 weeks of follow-up was needed to assess resource use beyond this period. Patients were excluded if they transferred out of their practice during the study period for any reason other than death. This was done to capture the full history of resource use and expenditure.

Two main cohorts of patients were sampled. The exposed cohort were adults with a SARS-CoV-2 infection confirmed by a reverse transcriptase polymerase chain reaction (RT-PCR) or lateral flow antigen test (see supplementary table 4 for included SNOMED-CT codes) and had not been hospitalised 14 days before or 42 days after infection (within 28 days of infection with a ±14-day grace period for clinical coding delays).^17^ Long COVID is underdiagnosed and poorly coded in primary care records and hence coded diagnoses of long COVID were not used to define the exposed cohort.^18^ The unexposed cohort consisted of propensity score-matched adults with no record of a positive RT-PCR or lateral flow antigen test for SARS-CoV-2 and no documented diagnoses of suspected or confirmed COVID-19, during the study period and had not been hospitalised during a matched time period. Within the exposed cohort, two subgroups were defined as those with a coded diagnosis of long COVID (DLC) and those reporting at least one of the recognised symptoms in the WHO diagnostic criteria for long COVID (SLC), 12 weeks after initial infection (Supplementary Tables 2 and 3).

### Propensity score matching

Propensity score matching (1:1) was used to closely match patients from the exposed and unexposed cohorts on several important confounding factors including age, sex, body mass index (BMI), smoking status, ethnic group, socioeconomic status (Index of Multiple Deprivation, IMD),^19^ index date, follow-up time from the index date, registered general practice, the number of primary care consultations in the 12 months prior to the index date (to account for informed presence bias),^20^ comorbidities and geographical region (Supplementary Table 1).

Propensity scores were derived from a logistic regression model of the probability of having a COVID-19 diagnosis as a function of 10 categories of covariates (Supplementary Table 1), with a calliper width of one-quarter of the standard deviation of the propensity score (0.04).^**Error! Reference source not found**.^ If matching is implemented properly, it can be assumed that the only systematic difference between the exposed and unexposed cohorts is the diagnosis of COVID-19, identifying the costs specifically caused by the disease.^22^ The matching performance was evaluated by comparing kernel density plots before and after matching to check the distribution of propensity scores and using the standardised differences between the groups for each variable, where a value greater than 0.1 was considered to indicate imbalance in baseline characteristics.

### Follow-up

The follow-up period was defined as the time between a patient’s index date (date of SARS-CoV-2 infection in the exposed cohort or matched time point in the unexposed cohort) and the patient’s study end date. This was defined as the earliest of the following time points: study end date (15^th^ April 2021), death date, or the last date of data collection from the practice contributing to the CPRD Aurum database.

### Outcomes and costing method

The primary outcome was the occurrence of a primary care consultation, defined as either a GP, nurse, or physiotherapy appointment. This included ten subcategories of healthcare professionals and four categories of consultation types (telephone appointments, surgery appointments, home visits, and telephone triage: see Supplementary Tables 5 and 6).

The costs of these consultations were estimated. Costs were assigned to the most specific cost data available. Unit costs for healthcare resources were taken from the Personal Social Services Research Unit’s (PSSRU) Unit Costs of Health and Social Care 2021, to account for inflation and variations in pricing over time, and to represent the cost perspective of the UK National Health Service (NHS).^23^ The hourly cost was available for each healthcare professional and the average consultation duration was used to calculate consultation costs.

Multiple consultations on the same day with the same healthcare professional were counted as a single consultation.^24^ Further details are provided in Supplementary Tables 7-9.

The incremental cost of primary care consultations between the exposed and unexposed cohorts was then estimated. The primary costing method was an ‘incremental cost’ approach. Bottom-up costing was adopted to estimate the healthcare use, with each patient’s resource utilisation calculated using individual-level data on consultations from CPRD Aurum.^26^ The healthcare utilisation was then multiplied by its respective unit cost and summed to obtain a patient’s total cost as follows:

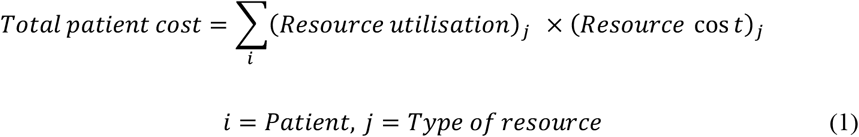

The incremental cost associated with long COVID was then obtained by subtracting the sum of each patient’s total healthcare cost between the matched groups, 12 weeks after the index date:

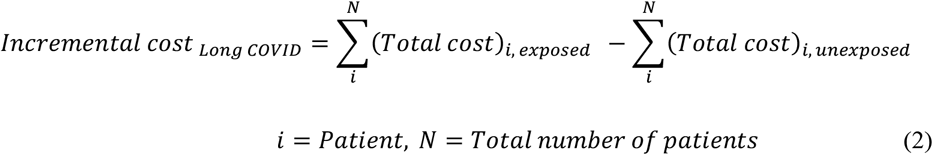

Usually, a cost of illness (COI) study includes a full year of data on all patients, as otherwise different follow-up times could be a major confounder. However, time since infection was included as a variable within the propensity score matching model to ensure similar follow-up time distributions in both cohorts (Supplementary Figure 1).

### Statistical Analysis

The analysis was split into three parts. The first estimated the incremental cost for primary care consultations associated with COVID-19 beyond 12 weeks from infection (i.e., costs associated with long COVID). The second part estimated the costs of these consultations across the UK population. The third part investigated the association between demographic and clinical risk factors and the cost of primary care consultations beyond 12 weeks from infection among those with a history of SARS CoV-2 infection.

In the first part of the analysis, the difference between the matched groups in total costs for primary care consultations was calculated within the matched follow-up period. Bootstrapped t-tests and analysis of variance (ANOVA) were used to compare means across the exposed and unexposed cohorts and the predefined subgroups. Bootstrapping means no distributional form was assumed and is standard practice for highly skewed cost data.^27^ A multivariable ordinary least squares (OLS) regression model was also used to assess the incremental cost while adjusting for relevant confounding factors. Although healthcare cost data is highly positively skewed, OLS assumptions are assumed not violated due to a large sample size.^28^ The proportion of consultation costs associated with each professional group (GP, nurse, and physiotherapist) and consultation type (telephone, in-person appointment, home visit, and triage) was also calculated.

A sensitivity analysis was conducted to assess the assumption that follow-up time does not confound the costs. The dataset was restricted to only patients who had at least six months of follow-up time from their index date. Only cost data from three to six months from the index date was included in the sensitivity analysis. Supplementary Figure 2 depicts a timeline showing the study dates and time periods of interest.

In the second part of the analysis, cumulative COVID-19 incidence estimates produced by the UK Office for National Statistics (ONS) in the COVID-19 Infection Survey, were used to estimate the national incremental costs attributed to primary care consultations for non-hospitalised patients with long COVID across the whole UK population.^29^ This was done by multiplying the population size by the cumulative incidence of COVID-19, as well as the 90% credible intervals. This provided the cumulative frequency of COVID-19. This was multiplied by the proportion of non-hospitalised patients with COVID-19 who had received at least one primary care consultation within the follow-up period in our study data. This value was then multiplied by the incremental cost estimated in the first part of the analysis to provide the national incremental cost of primary care consultations associated with long COVID. This was done for England, Wales, Scotland, and Northern Ireland, and for the total UK population. This was done separately using the estimated incremental costs from both the primary and sensitivity analyses. We assumed that the incremental costs would remain constant throughout the pandemic.

In the third part of the analysis, an econometric model was used to explore the cost predictors of primary care consultations in patients with a history of confirmed SARS-CoV-2 infection. We used a multivariable log ordinary least squares (OLS) regression model, which is suitable for positive and highly skewed healthcare cost data.^30, 31^ Log OLS models transform the dependent variable (individual patient cost) by the natural logarithm, which suppresses outlier values. Only patients who incurred a positive cost were included. This is because many patients had no consultations and thus zero cost, causing a skewed distribution of the dependent variable. The model included the DLC and SLC subgroups as covariates and adjusted for the same covariates used for the propensity score model (Supplementary Table 1). The covariates were checked for importance using a backward selection process with a threshold of p<0.1 to determine covariate inclusion. A p-value smaller than 0.05 was considered to indicate that a variable was statistically different from zero. Missing data was denoted by a missing category within the variable.

All statistical analyses were performed using STATA version 17 and R version 99.9.9.

## Results

### Study population

There were 472,173 patients in both the exposed and unexposed cohorts. The diagnosed long COVID (DLC) and symptomatic long COVID (SLC) subgroups consisted of 3,871 (0.8%) and 30,174 (6.4%) patients, respectively. 14% of the exposed cohort, 13% of the unexposed, 11% of the DLC subgroup, and 33% of the SLC subgroup had at least 6 months of follow-up data and were eligible for inclusion in the sensitivity analysis.

The kernel density plots (in Supplementary Figure 4) show that the matching was well-balanced because the density estimation lines for the two groups coincide. The standardised differences across covariates were less than 0.1 for all variables after matching (Supplementary Figure 5), which is further evidence of balanced matching. The matched groups were very similar in each of the baseline characteristics including age, sex, ethnic group, socioeconomic status, smoking status, BMI, the number of prior consultations, and a wide range of comorbidities (Table 1 and Supplementary Table 10). The mean age was 44 years, 55% were female, and 64% belonged to a white ethnic group. 22% were current smokers and just over 55% were overweight or obese.

**Table 1.** Baseline characteristics of the matched exposed and unexposed groups

| Variables | Unexposed<br>(n = 472,173)<br>n (%) | Exposed<br>(n = 472,173)<br>n (%) |
| --- | --- | --- |
| Age (mean (SD)) | 44.14 (16.92) | 44.16 (16.86) |
| Sex |  |  |
| Male | 210,848 (44.7) | 211,683 (44.8) |
| Female | 261,325 (55.3) | 260,490 (55.2) |
| Ethnicity |  |  |
| White | 300,873 (63.7) | 299,609 (63.5) |
| Asian | 59,720 (12.6) | 60,544 (12.8) |
| Black | 18,572 (3.9) | 18,598 (3.9) |
| Mixed | 9,410 (2.0) | 9,448 (2.0) |
| Other | 7,266 (1.5) | 7,208 (1.5) |
| Missing | 76,332 (16.2) | 76,766 (16.3) |
| IMD |  |  |
| 1 (Least deprived) | 74,314 (15.7) | 74,123 (15.7) |
| 2 | 76,964 (16.3) | 76,637 (16.2) |
| 3 | 80,474 (17.0) | 80,293 (17.0) |
| 4 | 96,097 (20.4) | 96,506 (20.4) |
| 5 (Most deprived) | 102,825 (21.8) | 103,331 (21.9) |
| Missing | 41,499 (8.8) | 41,283 (8.7) |
| Smoking Status |  |  |
| Current smoker | 104,696 (22.2) | 104,986 (22.2) |
| Ex-Smoker | 165,128 (35.0) | 163,759 (34.7) |
| Never smoked | 159,371 (33.8) | 159,655 (33.8) |
| Missing | 42,978 (9.1) | 43,773 (9.3) |
| BMI category |  |  |
| Normal weight | 143,346 (30.4) | 144,426 (30.6) |
| Underweight | 17,363 (3.7) | 17,483 (3.7) |
| Obese | 127,060 (26.9) | 125,086 (26.5) |
| Overweight | 139,589 (29.6) | 138,694 (29.4) |
| Missing | 44,815 (9.5) | 46,484 (9.8) |
| Number of consultations 3 to 12 months prior to index date (mean (SD)) |  |  |
| GP | 2.12 (3.68) | 2.15 (3.44) |
| Nurse | 0.50 (1.59) | 0.50 (1.49) |
| Physiotherapist | 0.01 (0.19) | 0.01 (0.19) |
| Surgery | 1.22 (2.24) | 1.20 (2.03) |
| Home visits | 0.03 (0.45) | 0.07 (0.79) |
| Telephone | 1.37 (2.73) | 1.38 (2.52) |
| Triage | 0.02 (0.30) | 0.02 (0.27) |

Notes: n: The number of patients in that category. %: Percentage of the group in the category. SD: Standard deviation.

Please insert Table 1 here.

### Incremental costs

The number of consultations and associated costs for patients 12 weeks from their index date for the period 15^th^ April 2020 to 15^th^ April 2021, stratified by the exposure status, is shown in Table 2. The numbers of primary care consultations were 209,620 (i.e., 0.44 per patient) in the unexposed cohort and 245,177 (i.e., 0.54 per patient) in the exposed cohort, respectively. Accordingly, patients in the exposed cohort had a 22.7% higher relative rate of consultations, in comparison to patients in the unexposed cohort. The total incremental cost of primary care consultations beyond 12 weeks from infection for the exposed cohort compared to the unexposed cohort was £2.44 per patient per year. Using OLS regression, the coefficient for belonging to the exposed cohort is interpreted as a £2.09 cost increase per exposed patient, supporting the main analysis (Supplementary Table 11). Patients in the DLC and SLC subgroups had consultation rates over 3 and 6 times greater than the unexposed cohort, respectively. This is an incremental cost of £30.52 and £57.56 per patient.

**Table 2.**
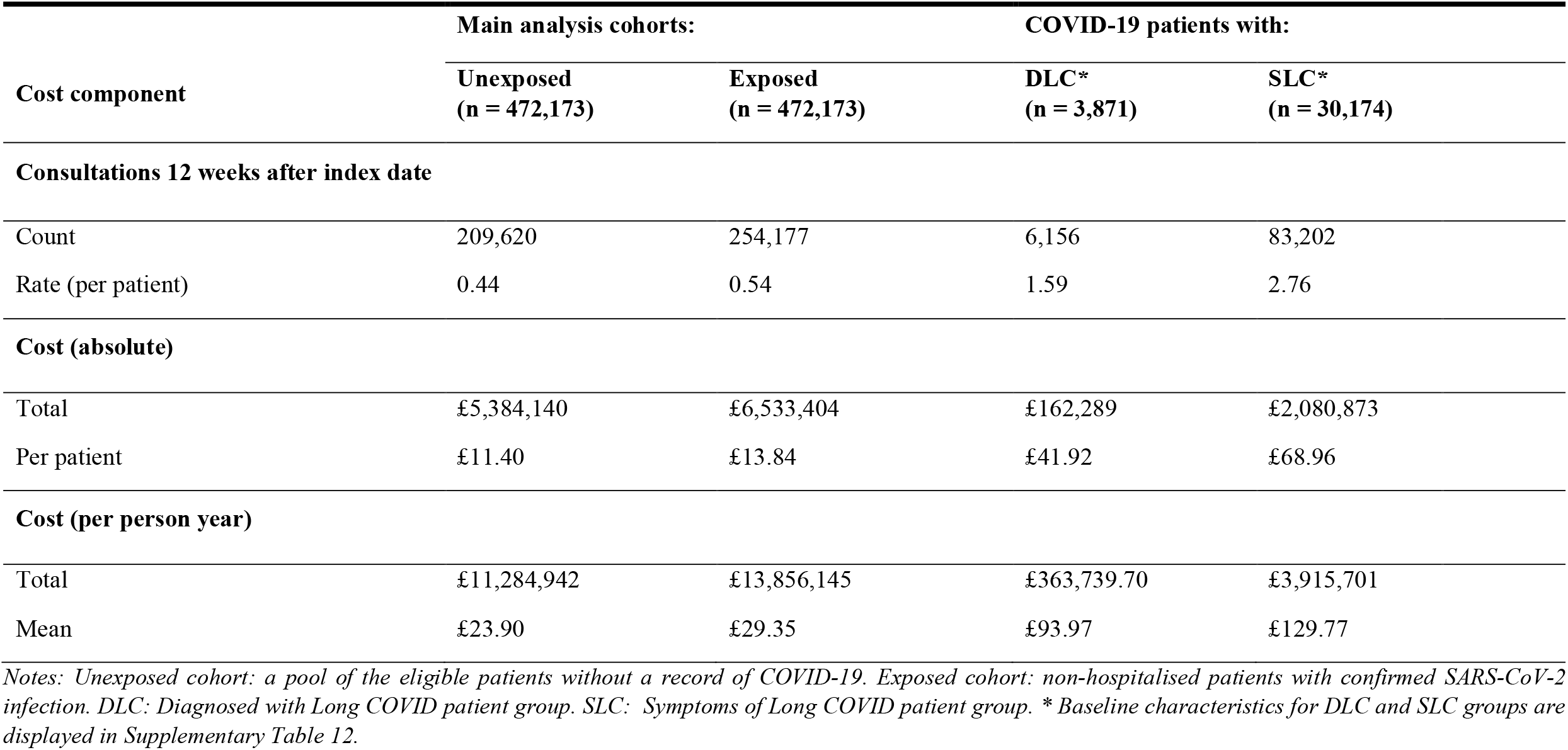
Estimates of the annual primary care resource use and costs associated with Long COVID between 15^th^ April 2020 to 15^th^ April 2021

Please insert Table 2 here.

GP consultations were the largest contributor to total costs for each of the exposure groups, representing over 85% of costs (Figure 1). GP consultations made up proportionately more of the total cost for the exposed and DLC and SLC subgroups than the unexposed cohort. The average cost per patient was higher for all COVID-19 related groups in comparison to patients in the unexposed cohort. Across each type of healthcare professional, the SLC subgroup was the most expensive per patient.

**Figure 1.**
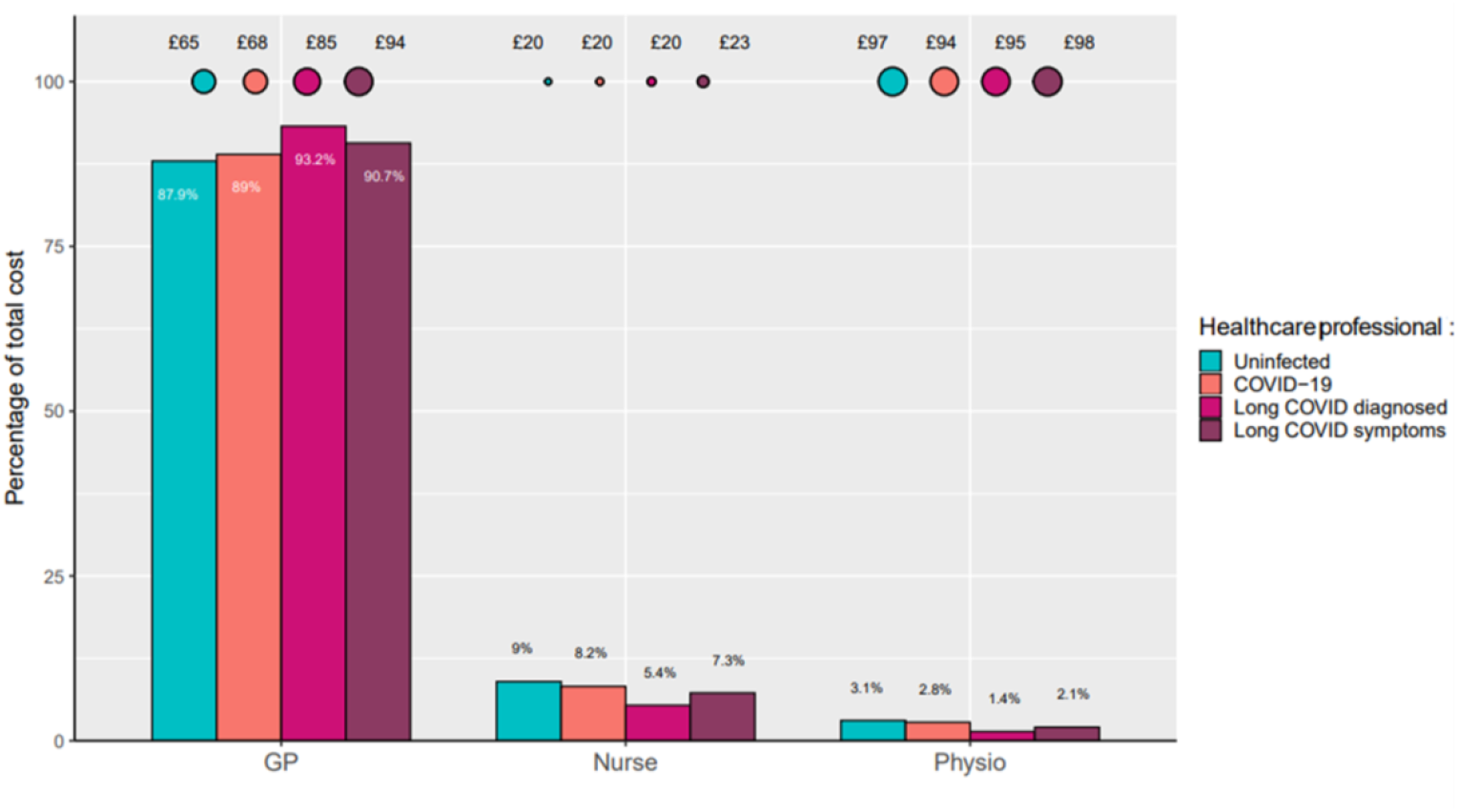
Bubble plot showing the average cost of each healthcare professional per patient who had a consultation for 15^th^ April 2020 to 15^th^ April 2021. Bar chart to show the percentage makeup of each group’s total costs by healthcare professional.

Please insert Figure 1 here.

For all groups, telephone consultations were the biggest contributor to total costs (over 60%) and were highest in the DLC and SLC subgroups (Figure 2). By contrast, the burden of in-person consultations on total costs was greatest in the unexposed cohort. Home visits made up a relatively large amount of costs for the exposed cohort and SLC subgroup, in comparison to the other groups. The average incremental costs of home visits for these groups were £19 and £35 higher than for patients in the unexposed cohort, respectively.

**Figure 2.**
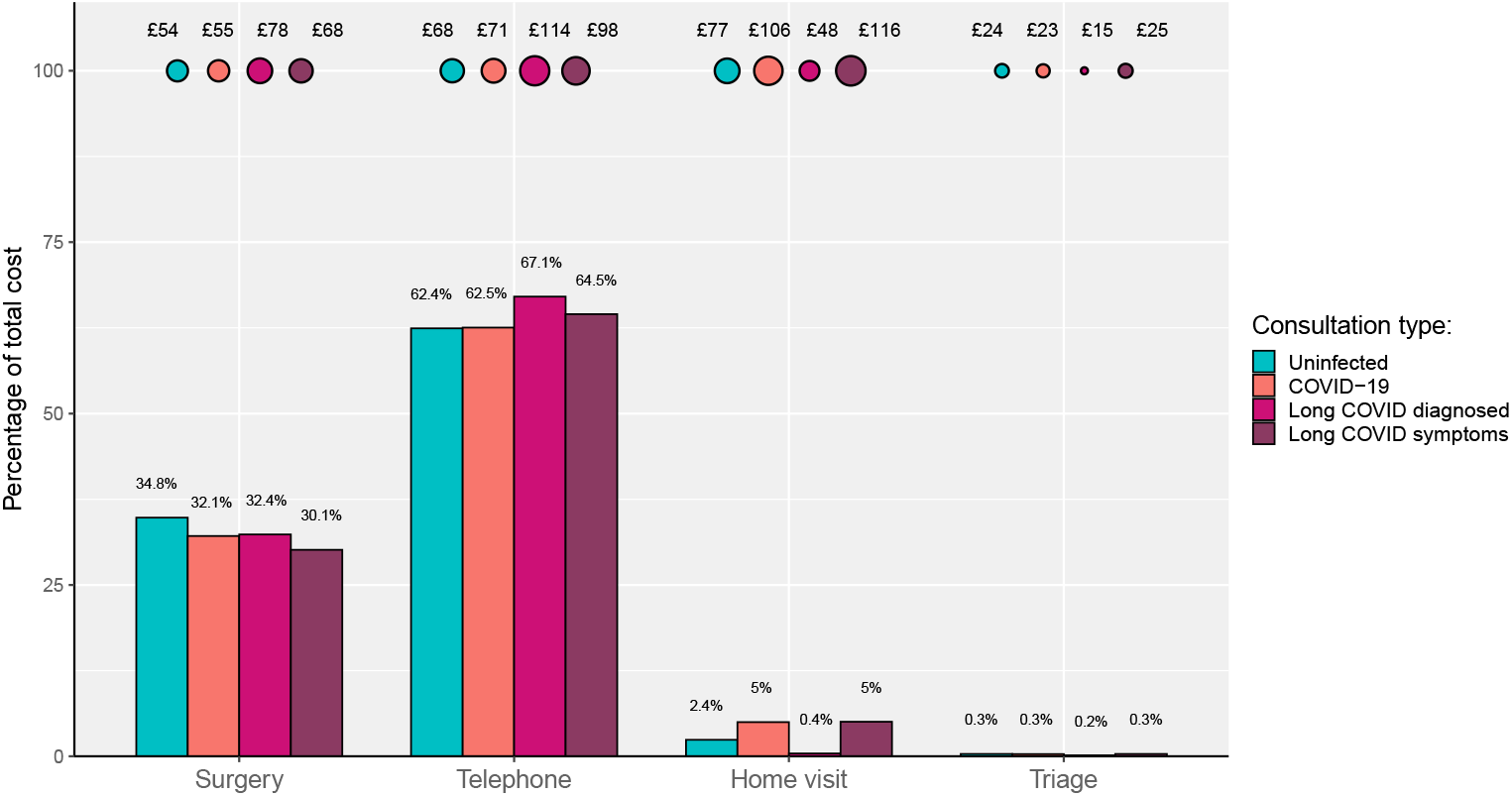
Bubble plot to show the average cost of each consultation type per patient (who had a consultation) for 15^th^ April 2020 and 15^th^ April 2021. Bar chart to show the percentage makeup of total costs by consultation type.

Please insert Figure 2 here.

The results of the sensitivity analysis are presented in Supplementary Table 14 and Supplementary Figure 7, which relate to costs among participants who had a minimum of six months of follow-up from their index date. These followed a similar trend to the cost per person year estimates in the main analysis although the overall incremental costs were higher. The exposed cohort was more costly than the unexposed cohort, with an incremental cost of £5.72 per patient. Patients in the DLC subgroup were the most expensive, followed by the SLC subgroup, with incremental costs per patient of £68.55 and £46.98, respectively when compared to the unexposed cohort.

### Estimation of national incremental costs

Using estimates of the cumulative incidence of COVID-19 in the ONS COVID-19 Infection Survey and applying an average incremental cost of £2.44 per patient, we estimate the additional primary care consultations costs in the UK associated with long COVID to total £23,382,452 (90% credible interval £21,378,567 to £25,526,052) (Table 3). When applying an average incremental cost of £5.72, based on the sensitivity analysis, we estimate these costs to be £54,814,601 (90% credible interval £50,116,967 to £59,839,762).

**Table 3.**
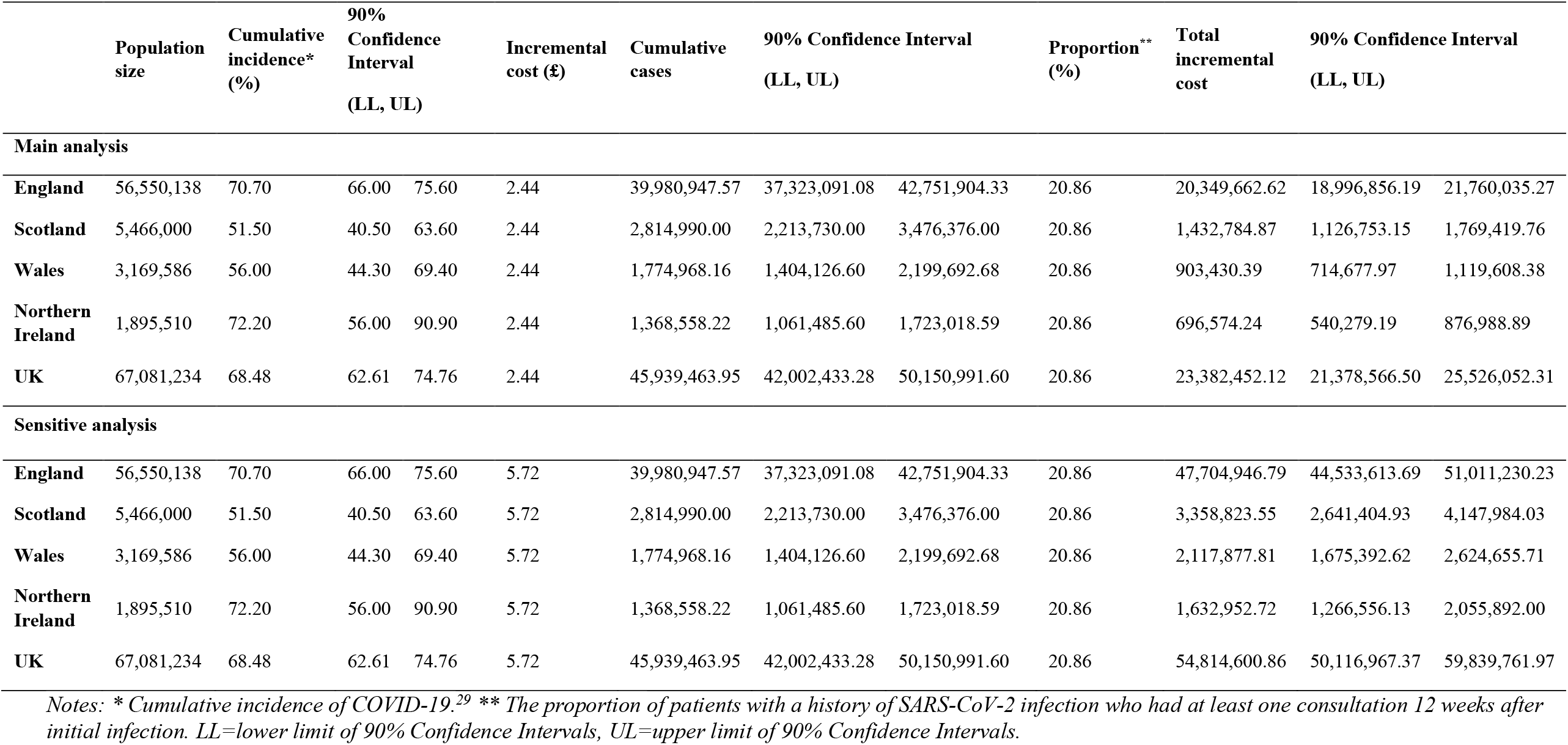
Primary care consultations costs in the UK

Please insert Table 3 here.

### Risk factor analysis

The results of the log OLS regression model are presented in Table 4. The coefficients are reported in exponential form. These are interpreted as the percentage change in total cost due to a one unit increase for continuous variables, or the presence of a categorical variable. The coefficients for having a diagnosis of long COVID or having symptoms of long COVID, were both statistically significant and corresponded to a 43% and 44% increase in primary care consultation costs in comparison to patients with a history of COVID-19 but no record of a Long COVID diagnosis or associated symptoms.

**Table 4.** Regression estimates for the log ordinary least squares (OLS) model on primary care consultation costs of patients with Covid-19 at least 12 weeks after infection

| Total healthcare cost | (Exp) Coef. * | 95% Confidence Intervals |  | p-value |
| --- | --- | --- | --- | --- |
| Exposure status |  |  |  |  |
| COVID-19 (Reference group) |  |  |  |  |
| Long COVID diagnosis | 1.43 | 1.34 | 1.52 | <0.001 |
| Symptoms of long COVID | 1.44 | 1.41 | 1.48 | <0.001 |
| Age (at index date) |  |  |  |  |
| 18-29 (Reference group) |  |  |  |  |
| 30-39 | 1.03 | 1.00 | 1.07 | 0.04 |
| 40-49 | 1.05 | 1.02 | 1.09 | <0.001 |
| 50-59 | 1.07 | 1.03 | 1.11 | <0.001 |
| 60-69 | 1.15 | 1.10 | 1.20 | <0.001 |
| 70-79 | 1.37 | 1.30 | 1.43 | <0.001 |
| ≥80 | 1.49 | 1.42 | 1.57 | <0.001 |
| Sex |  |  |  |  |
| Male (Reference group) |  |  |  |  |
| Female | 1.04 | 1.01 | 1.06 | 0.01 |
| Ethnic group |  |  |  |  |
| White (Reference group) |  |  |  |  |
| Black | 0.94 | 0.88 | 0.99 | 0.03 |
| Other | 1.06 | 0.96 | 1.18 | 0.25 |
| Asian | 1.00 | 0.97 | 1.03 | 0.97 |
| Mixed | 0.99 | 0.93 | 1.05 | 0.64 |
| Ethnicity Missing | 0.99 | 0.96 | 1.03 | 0.76 |
| Socioeconomic status (IMD) |  |  |  |  |
| 1 (Least deprived) (Reference group) |  |  |  |  |
| 2 | 1.08 | 1.04 | 1.13 | <0.001 |
| 3 | 1.07 | 1.02 | 1.11 | <0.001 |
| 4 | 1.01 | 0.96 | 1.06 | 0.68 |
| 5 (Most deprived) | 1.05 | 1.01 | 1.10 | 0.01 |
| IMD Missing | 1.00 | 0.94 | 1.06 | 0.92 |
| Smoking status |  |  |  |  |
| Never Smoked (Reference group) |  |  |  |  |
| Ex-Smoker | 1.01 | 0.98 | 1.04 | 0.65 |
| Current Smoker | 1.00 | 0.96 | 1.03 | 0.83 |
| Smoker Missing | 1.00 | 0.96 | 1.05 | 0.88 |
| BMI categories |  |  |  |  |
| Normal weight (Reference group) |  |  |  |  |
| Underweight | 1.02 | 0.95 | 1.11 | 0.55 |
| Overweight | 0.98 | 0.95 | 1.01 | 0.20 |
| Obese | 1.04 | 1.01 | 1.07 | 0.02 |
| BMI Missing | 1.09 | 1.03 | 1.16 | 0.01 |
| Charlson Comorbidity Index | 1.01 | 1.01 | 1.02 | <0.001 |
| GP consultations prior** | 1.03 | 1.03 | 1.04 | <0.001 |
| Nurse consultations prior | 1.01 | 1.01 | 1.02 | <0.001 |
| Physiotherapist consultations prior | 1.04 | 1.01 | 1.07 | <0.001 |
| Weeks since index date | 1.04 | 1.04 | 1.04 | <0.001 |

Notes: \*Difference in cost from a one-unit change or in comparison to the reference group – e.g., 1.43 refers to a 43% relative increase in costs due to a one-unit increase or compared to the reference group. These have been adjusted for the covariates in the table in addition to geographic region.
\*\*Prior consultations are the sum of the number of consultations a patient had in the 3 to 12 months prior to their index date, with the healthcare professional specified.

Please insert Table 4 here.

Older age (49% relative increase in costs in those aged 80 years or older compared to those aged 18 to 29 years), female sex (4% relative increase in costs compared to males), obesity (4% relative increase in costs compared to those of normal weight), comorbidities and frequency of prior consultations were all associated with an increase in the cost of primary care consultations. Those from black ethnic groups had a 6% reduced cost compared to those from white ethnic groups, although no significant differences were seen between white ethnic groups and other minority ethnic groups. While patients from the second, third, and fifth most socioeconomically deprived quintiles had higher costs than those from the least deprived quintile, the differences in these costs did not follow a clear gradient.

## Discussion

### Main findings

In this study of over 470,000 non-hospitalised patients with a history of SARS-CoV-2 infection and closely matched individuals with no history of COVID-19, we found that those with a history of infection cost primary care services on average an additional £2.44 per patient for primary care consultations at least 12 weeks after infection. However, this incremental cost could be as high as £5.72 per patient. The incremental costs were significantly higher for those diagnosed with long COVID (£30.52) and those documented as reporting associated symptoms (£57.56). Most of these additional costs were from GP telephone consultations. We estimate that the national costs for primary care consultations to support people with long COVID in the UK are approximately £23 million but may approach £60 million.

Among those with a history of SARS-CoV-2 infection, higher consultation costs were associated with having a diagnosis or reporting symptoms of long COVID, older age, being female, and obesity. While the most affluent socioeconomic quintile had lower costs than those from more deprived socioeconomic groups, there was no clear socioeconomic gradient in incremental costs. By contrast, those from black ethnic groups incurred lower costs than those from white ethnic groups, while there was no difference with other ethnic groups. This highlights a potential health inequality, especially given the poorer outcomes (e.g., more hospital admissions, higher mortality rate of death) following COVID-19 among individuals from black ethnic minority groups.^32, **Error! Reference source not found**.^

### Relationship to other studies

Whittaker et al. (2021) used data from the CPRD Aurum database to assess consultation rates for patients with COVID-19.^**Error! Reference source not found**.^ They reported that they had significantly higher GP consultation rates, which led to an 18% increase in healthcare utilisation post-infection compared to the 12 months prior. Furthermore, patients with COVID-19 continued to display higher GP consultation rates even four weeks after infection. We further show that this trend continued beyond 12 weeks after SARS-CoV-2 infection and have estimated associated consultation costs.

Another population-based retrospective cohort study by Koumpias et al. (2022) assessed the healthcare use and costs of over 250,000 patients with a history of COVID-19 using administrative claims data in the United States from March to September 2020.^34^ This study included a wide range of healthcare resources, including use of telemedicine, urgent care, and inpatient services. They found that monthly costs increased significantly following COVID-19 compared to prior to infection, with additional costs persisting beyond five months, particularly among adults aged older than 45 years. The study however did not have a contemporary control group and did not delineate between primary and secondary care services.

Calderón-Moreno et al. (2022) investigated the primary care costs associated with COVID-19.^35^ They assessed 6,286 patients with a diagnosis of COVID-19 in a primary care setting from the Spanish region Aragon, estimating an average illness-associated cost of €729.79 per patient. The costing approach was unclear and there are difficulties in comparing healthcare costs between countries, but the study highlighted the significant economic burden of the illness.^35^ The authors found that the complications arising from COVID-19, such as respiratory, cardiovascular, and haematological disorders, caused substantial further cost increases. However, the study did not specifically comment on the costs associated with long COVID.

There is also broader literature on the impact of COVID-19 on the utilisation of primary care resources. For many patients, especially those with less severe illnesses, the pandemic led to a reduction in overall healthcare use, but an increase in the number of non-face-to-face consultations.^36^ We similarly found that the increased cost of primary consultations associated with long COVID were driven by an increase in telephone consultations.

### Strengths and limitations

A key strength of the study was that the costs associated with long COVID could be isolated by implementing an incremental cost approach using a highly matched comparison group with no prior history of suspected or confirmed COVID-19. The matching algorithm was comprehensive and included many relevant variables that are associated with healthcare costs and was successful in achieving a close balance in baseline demographic and clinical characteristics between the exposed and unexposed cohorts. This was fundamental to the inferences being made, as except from unobservable factors, the only key difference between the cohorts were the record of SARS-CoV-2 infection.^22^ Another strength was the large sample size, which included 944,346 patients. This helped to provide two closely matched cohorts, results that are likely to be representative of the UK population, and high statistical power for our analyses.^14, 37^

This study is subject to several limitations. A key limitation of the data source was the lack of diagnosis of long COVID in primary care records.^18^ However, in our analysis we only incorporated costs for consultations that occurred at least 12 weeks after confirmation of SARS-CoV-2 infection (or matched time point for the unexposed cohort). We inferred that any differences in consultation costs beyond this time point were likely to be attributable to the longer-term effects of COVID-19 or long COVID since both cohorts were very closely matched in their demographic and clinical characteristics except for SARS-CoV-2 infection. We also assessed costs in two subgroups within the exposed cohort that had either diagnosed long COVID or had records of symptoms that overlapped with those associated with long COVID. As long COVID diagnosis improves over time, we would expect clinical coding of this diagnosis to improve, which should assist with future economic studies.

Second, the duration of consultations is not well recorded, and costs based on duration were unable to be calculated. Unit costs of primary care consultations were therefore obtained from the PSSRU’s Unit Costs of Health and Social Care 2021 for primary care consultations, which assumed a standard duration of consultation for each patient. However, there is evidence suggesting that the duration of primary care consultations can vary by many factors, such as doctor-related factors (e.g., gender, experience) and patient-related characteristics (e.g., number of conditions, socioeconomic status).^38^ Thus, actual costs of primary care consultations may vary from the estimated costs in this study. Furthermore, when estimating the national costs from consultations associated with long Covid, we assumed that incremental costs would remain constant over the course of the pandemic, which may not necessarily be true as access to primary changed during this period.

Third, although we used propensity score matching to reduce the risk of confounding, there may still be residual confounding accounting for differences in consultation rates between the exposed and unexposed cohorts. For example, we did not have access to data on occupational status, which might be associated with both the risk of COVID-19 infection as well as consultation rates. Furthermore, we were unable to control for SARS CoV-2 vaccination status in propensity score matching and regression modelling, although a relatively small proportion of the population had been vaccinated during that period of the pandemic.

However, we anticipate that residual confounding would be limited in our results, given the wide range of demographic and clinical covariates considered.

Another limitation is the potential misclassification of individuals in the unexposed cohort. Community based testing for SARS-CoV-2 was relatively limited in the UK during the first wave of the COVID-19 pandemic.^39^ Some members of the unexposed cohort may have had COVID-19 but not been formally tested. We attempted to limit this by excluding patients from the unexposed cohort if they had a record of either suspected or confirmed COVID-19, even in the absence of any confirmatory testing. However, this is unlikely to have completely removed this source of misclassification bias and is therefore likely to have reduced our effect size, and therefore underestimated the true incremental cost of long COVID from primary care consultations.

### Implications for practice, policy and research

Our analysis suggests that the cost of supporting non-hospitalised adults with long COVID in primary care is likely to be substantial, even when only considering consultation costs. This is at a time of exceptional pressure on health services, including primary care in the UK and worldwide. Primary care services in the UK are likely to need in the order of £20-£60 million to support primary care consultations in patients with long COVID, the majority of which will be needed for remote GP consultations. The scale of these costs is likely to be similar in other comparable health settings. It should be noted that some non-hospitalised patients with COVID-19 are likely require secondary care referral, which has further cost implications not considered in the current study. Overall, this will require substantial investment globally to ensure that primary care services are adequately resourced to provide the complexity of care needed to support non-hospitalised patients with ongoing symptoms and care needs. Training allied healthcare professionals to support this care, with implementation of guidelines for long COVID diagnosis and care,^40^ could potentially help to reduce these costs.

Our analysis also suggests that in addition to those with an established diagnosis of long COVID, those with a history of SARS-CoV-2 infection without a formal diagnosis of long COVID but reporting relevant symptoms to primary care clinicians are likely to place significant additional costs for primary care consultations. Furthermore, certain population subgroups amongst those with a history of SARS-CoV-2 infection are likely to incur increased costs, such as the elderly, females, and those with obesity. Additionally, those from black ethnic groups may be underusing primary care services for long COVID symptoms, representing a potential health inequity. These factors should be considered by health service commissioners, managers and providers when designing and resourcing long COVID services in primary care.

This study provides a foundation in methods and cost estimates for future cost analyses and economic evaluations on long COVID. Future research should focus on updating this analysis to capture longer-term patient data and costs, evaluate the impact of long COVID on prescription drug costs, assess secondary care costs, assess out-of-pocket costs, and explore methods to better capture costs specifically attributable to long COVID.

## Conclusion

The support of non-hospitalised individuals with long COVID in primary care is likely to be substantial, requiring significant healthcare investment and planning. This particularly applies to patients who have been formally diagnosed with long COVID, those without a long COVID diagnosis but with a history of SARS-CoV-2 infection and reporting related symptoms, the elderly, females, and those with obesity. Inequalities in access to primary care services for long COVID support require further exploration and need to be addressed.

## Data Availability

Following protocol approval from the MHRA Independent Scientific Advisory Committee, access to anonymised patient data from the Clinical Practice Research Datalink (CPRD) is authorised based on the data sharing agreement with specific terms and conditions of usage. Therefore, the dataset used in this study is not available to the public. Requests for access to the data used for this study will need to be directed to CPRD.

## Contribution statement

JT undertook the analysis and drafted the report with input from all co-authors. SH, LJ, DZ, and KN conceived the idea for the study and study design. LJ and DZ provided oversight and advised on the health economic and econometric analyses. PM, TM, MC, TW reviewed the results and provided feedback on the draft manuscript. SH provided clinical, public health and epidemiological oversight of the study. KN, AS, and KG provided data science and epidemiological expertise and support. NG provided input on health economic methods and analyses and revisions of the manuscript. KM is a public partner to the TLC study and reviewed the manuscript. All authors reviewed and approved the final version.

## Ethics approval

CPRD supplies anonymised UK health data for public health research sponsored by the National Institute for Health Research (NIHR) and the UK Medicines and Healthcare products Regulatory Agency (MHRA). The UK’s Health Research Authority Research Ethics Committee provides ethics approval to CPRD annually. Therefore, no additional ethics approval is required for observational studies using CPRD Aurum data for public health research, subject to individual research protocols meeting CPRD data governance requirements. Access to the CPRD database in this study obtained the protocol approval from the CPRD Independent Scientific Advisory Committee (Reference ID: 22_001855).

## Patient and public involvement statement

The TLC Study has had extensive patient and public involvement through its PPI group. This manuscript has been reviewed and received critical feedback and was co-authored by a PPI group member (KM).

## Dissemination to participants and related patient and public communities

The findings of this study have been presented to general practitioners in the West Midlands. The UK Department of Health and Social Care has been informed of the findings. The findings will be presented at national and international conferences, invited talks, workshops or webinars. We will distribute the article to both clinicians and long covid support groups. We will also distribute findings on social media and will be reported in a plain language summary on the Therapies for Long COVID Study website (www.birmingham.ac.uk/research/applied-health/research/long-covid/index.aspx).

