## Supplementary Material for "The cost of primary care consultations associated with long COVID in non-hospitalised adults: a retrospective cohort study using UK primary care data"

**Table of Contents**

**Supplementary Table 1.** Table of explanatory variables included in the study

| **Variable** | **Description** | **Specification** | **Models** |
| --- | --- | --- | --- |
| Age | Age in years at index date | Continuous | * f |
| Age group | Age categorised into 8 groups:  18-29, 30-49, 50-59, 60-69, 70-79, 80+ | Derived categorical | g |
| Sex | Male or Female | Binary | * g f |
| BMI category | Underweight <18.5 kg/m2,  Normal weight 18.5-24 kg/m2  Overweight 25-29 kg/m  Obese ≥30 kg/m2  BMI Missing | Derived categorical | * g f |
| Index of Multiple Deprivation | Measure of socio-economic status from 1 to 5:  1 – Least deprived  5 – Most deprived  IMD missing | Categorial | * g f |
| Smoking status | Never smoked  Ex-smoker  Current smoker  Smoker Missing | Categorial | * g f |
| Ethnicity | White  Black  Mixed  Asian  Other  Ethnicity Missing | Categorical | * g f |
| Time since index | Number of weeks from index date to patient’s end of study period | Derived continuous | * f |
| Practice ID | Indicates which practice a patient belongs to. | Categorial | * |
| Prior healthcare utilisation | Number of consultations had with a healthcare professional in the 3 to 12 months prior to a patient’s index date:  GP  Nurse  Physiotherapist | Continuous | * f |

**Supplementary Table 1.** Table of explanatory variables included in the study (Continued)

| **Variable** | **Description** | **Specification** | **Models** |
| --- | --- | --- | --- |
| 87 Comorbidities | Chronic health conditions, one category with patient numbers ≤5 was suppressed in accordance with CPRD guidelines for data protection. The list of comorbidities can be found in Supplementary Table 11. | Binary | * |
| Charlson Comorbidity Index  (CCI) | Using a CCI R package, CCI was calculated for each patient. 17 comorbidities are included and are given different scores dependant on the range a patient has. | Derived continuous | f |
| Region | 11 regions from England and Northern Ireland:  Northeast, North West, East Midlands, West Midlands, South East, South West, East of England, South Central, London, Northern Ireland | Categorial | g f |

*Note: * Included in propensity score matching. g: Included in stratification of costs. f: Included in the regression specification.*

**Supplementary Table 2.** Study group definitions and titles

|  | **Main groups** | | **Sub-groups**  (Patients with a diagnosis of COVID-19…) | |
| --- | --- | --- | --- | --- |
| **Group title** | **Unexposed group** | **Exposed group** | **Diagnosed Long COVID (DLC) group** | **Symptoms of Long COVID (SLC) group** |
| **Definition** | Patients without a diagnosis of COVID-19 | Patients with a diagnosis of COVID-19 | … and a diagnosis of Long COVID | … and show at least one of the WHO symptoms of Long COVID after 12 weeks post infection |

**Supplementary Table 3.** Table of Long COVID symptoms classified by WHO Symptoms must be shown by a COVID-19 patient 12 weeks after their index date

| **Symptom** | **SNOMED CT Code** |
| --- | --- |
| Anosmia | 44169009 |
| Fatigue | 84229001 |
| Fever | 386661006 |
| Palpitations | 80313002 |
| Blurred vision/diplopia | 24982008 |
| Chest pain | 29857009 |
| Shortness of breath | 267036007 |
| Paraesthesia | 91019004 |
| Diarrhoea | 62315008 |
| Headache | 25064002 |
| Allergies | 370860007 |
| Insomnia | 193462001 |
| Constipation | 14760008 |
| Joint pain | 57676002 |
| Difficulty thinking | 285257002 |
| Abdominal pain | 21522001 |
| Gastritis | 4556007 |
| Dizziness | 404640003 |
| Menorrhagia | 386692008 |
| Hyperacusis | 25289003 |
| Muscle pain | 68962001 |
| Amnesia | 48167000 |
| Muscle cramping | 55300003 |
| Gastric reflux | 225587003 |
| Depression | 255339005 |
| Dysgeusia | 271801002 |
| Anxiety | 48694002 |
| Menstrual changes | 80182007 |
| Pre-menstrual syndrome | 289896006 |
| Excessive sleep | 77692006 |
| Hearing loss | 15188001 |
| Post exert fatigue | 444042007 |

**Supplementary Table 4.** Table of all descriptions of COVID-19 and relevant SNOMED CT codes used to define whether a patient had COVID-19

| **Description** | **SNOMED CT Code** |
| --- | --- |
| Detection of 2019 novel coronavirus using polymerase chain reaction technique | 1240511000000100 |
| Detection of 2019-nCoV (novel coronavirus) using polymerase chain reaction technique | 1240511000000100 |
| Detection of SARS-CoV-2 (severe acute respiratory syndrome coronavirus 2) using polymerase chain reaction technique | 1240511000000100 |
| Detection of Wuhan 2019-nCoV (novel coronavirus) using polymerase chain reaction technique | 1240511000000100 |
| Wuhan 2019-nCoV (novel coronavirus) detected | 1240581000000100 |
| 2019-nCoV (novel coronavirus) detected | 1240581000000100 |
| 2019 novel coronavirus detected | 1240581000000100 |
| SARS-CoV-2 (severe acute respiratory syndrome coronavirus 2) detected | 1240581000000100 |
| SARS-CoV-2 (severe acute respiratory syndrome coronavirus 2) detection result positive | 1240581000000100 |
| SARS-CoV-2 (severe acute respiratory syndrome coronavirus 2) RNA (ribonucleic acid) detection result positive | 1240581000000100 |
| COVID-19 detected | 1240581000000100 |
| COVID-19 confirmed by laboratory test | 1300721000000100 |
| SARS-CoV-2 (severe acute respiratory syndrome coronavirus 2) RNA (ribonucleic acid) qualitative existence in specimen | 1321301000000100 |
| SARS-CoV-2 (severe acute respiratory syndrome coronavirus 2) antigen detection result positive | 1322781000000100 |
| 2019-nCoV (novel coronavirus) antigen detection result positive | 1322781000000100 |
| SARS-CoV-2 (severe acute respiratory syndrome coronavirus 2) RNA (ribonucleic acid) detection result positive | 1324601000000100 |
| 2019-nCoV (novel coronavirus) ribonucleic acid detected | 1324601000000100 |
| SARS-CoV-2 (severe acute respiratory syndrome coronavirus 2) RNA (ribonucleic acid) detection result positive at the limit of detection | 1324881000000100 |
| 2019-nCoV (novel coronavirus) detection result positive at the limit of detection | 1324881000000100 |
| Detection of SARS-CoV-2 (severe acute respiratory syndrome coronavirus 2) antigen | 871553007 |
| Detection of ribonucleic acid of 2019 novel coronavirus in nasopharyngeal swab | 871556004 |
| Detection of RNA (ribonucleic acid) of SARS-CoV-2 (severe acute respiratory syndrome coronavirus 2) in nasopharyngeal swab | 871556004 |
| Detection of RNA (ribonucleic acid) of SARS-CoV-2 (severe acute respiratory syndrome coronavirus 2) in oropharyngeal swab | 871557008 |
| Detection of RNA (ribonucleic acid) of SARS-CoV-2 (severe acute respiratory syndrome coronavirus 2) using polymerase chain reaction | 871560001 |
| Detection of ribonucleic acid of COVID-19 using polymerase chain reaction | 871560001 |

**Supplementary Table 5.** Table of the CPRD Aurum codes and descriptions of healthcare professionals included in the study, grouped by the healthcare professional category

| **CPRD Aurum codes: jobcatid** | **Description** |
| --- | --- |
| **GP** |  |
| 4 | General Medical Practitioner |
| 5 | Salaried General Practitioner |
| 15 | Associate Practitioner - General Practitioner |
| 24 | GP Registrar |
| 31 | Sessional GP |
| 181 | Locum GP |
| 183 | Assistant GP |
| **General Nurse** |  |
| 9 | Community Nurse |
| 48 | Enrolled Nurse |
| 52 | Community Mental Health Nurse |
| 61 | Sister/Charge Nurse |
| **Staff Nurse** |  |
| 47 | Staff Nurse |
| 59 | Student Practice Nurse |
| 115 | Student District Nurse |
| 169 | Student Community Mental Health Nurse |
| 191 | Student Occupational Health Nurse |
| **Specialist Nurse** |  |
| 27 | Specialist Nurse Practitioner |
| 55 | Associate Practitioner - Nurse |
| **Nurse consultant** |  |
| 33 | Nurse Consultant |
| **Nurse Manager** |  |
| 60 | Nurse Manager |
| **Physiotherapist Consultant** |  |
| 106 | Physiotherapist Consultant |
| **Physiotherapist Specialist Practitioner** |  |
| 80 | Physiotherapist Specialist Practitioner |
| **Physiotherapist Manager** |  |
| 136 | Physiotherapist Manager |
| **Physiotherapist** |  |
| 34 | Physiotherapist |
| 77 | Student Physiotherapist |

**Supplementary Table 6.** Table of the CPRD Aurum codes and descriptions of consultation types included in the study, grouped by the consultation type category

| **CPRD Aurum codes: Consultation Source Id** | **Description** |
| --- | --- |
| **Surgery** |  |
| 751 | G.P. Evening Surgery |
| 4221 | Surgery consultation |
| 3509 | Nurse Surgery |
| 10310 | Surgery Clinic |
| 5266 | GP Surgery |
| 2706 | GP Surgery |
| 4231 | GP Surgery |
| 754 | GP Surgery |
| 7632 | GP Surgery |
| 7659 | GP Surgery |
| 7732 | GP Surgery |
| 9361 | Surgery or Clinic |
| 2711 | G.P Surgery (Pm) |
| 4215 | Surgery Attendance |
| 1865 | Branch Surgery |
| 2719 | G.P. Morning Surgery |
| 3505 | Nurse Practitioner Surgery |
| 6963 | Walk-In Surgery |
| 426 | Contact method: G.P.Surgery |
| 2580 | Emergency Gp Surgery |
| 5252 | G.P.Surgery Urgent Consultation |
| 10303 | Surgery |
| 9551 | Unbooked Clinic |
| 3499 | Nurse Assessment Clinic |
| 6140 | P.Nurse Clinic |
| 364 | Clinic NHS |
| 366 | Clinic note |
| 3346 | Minor Operations Clinic |
| 3503 | Nurse Minor Illness Clinic |
| 5093 | Emergency Nurse Clinic |
| 6236 | Practice Nurse Clinic |
| 7240 | Clinic Premises |
| 397 | Community Clinic |
| 10310 | Surgery Clinic |
| 4773 | Clinic |
| 5952 | Nurse's Treatment Room Clinic |
| 3347 | Minor Ops Clinic |

**Supplementary Table 6.** Table of the CPRD Aurum codes and descriptions of consultation types included in the study, grouped by the consultation type category **(Continued)**

| **CPRD Aurum codes: Consultation Source Id** | **Description** |
| --- | --- |
| 3501 | Nurse Flu Clinic |
| 3516 | Nurses' Flu Clinic |
| 7395 | Diabetic Clinic |
| 8812 | Out Of Hours Visit |
| 3631 | Out Of Hours Night Visit |
| 3627 | Out Of Hours Gp Visit |
| 5869 | Night Visit |
| 6087 | Out of hours, Practice |
| 16305 | Out of Hours |
| 6034 | Out of Hours |
| 6052 | Out of Hours |
| 6056 | Out of Hours |
| 6062 | Out of Hours |
| 6063 | Out of Hours |
| 8794 | Out of Hours |
| 8795 | Out of Hours |
| 8796 | Out of Hours |
| 3617 | Out of Hours |
| 3618 | Out of Hours |
| 8503 | Out of Hours |
| 8504 | Out of Hours |
| 8505 | Out of Hours |
| 8506 | Out of Hours |
| 6064 | Out of Hours |
| 6065 | Out of Hours |
| 6066 | Out of Hours |
| 3614 | Out of Hours |
| 3615 | Out of Hours |
| 3616 | Out of Hours |
| 8799 | Out of hours consultation at surgery |
| 3632 | Out Of Hours Other Practice |
| 9907 | Practice Nurse |
| 2810 | GP Practice |
| 5875 | Night visit, practice |
| 6236 | Practice Nurse Clinic |
| 9203 | Seen by Practice Nurse |
| 745 | G P Consultation |
| 421 | Consultation |

**Supplementary Table 6.** Table of the CPRD Aurum codes and descriptions of consultation types included in the study, grouped by the consultation type category **(Continued)**

| **CPRD Aurum codes: Consultation Source Id** | **Description** |
| --- | --- |
| 3626 | Out Of Hours GP Service |
| 9633 | Walk In Centre |
| 4523 | Walk-in clinic |
| 6963 | Walk-In Surgery |
| 5149 | Face to face consultation |
| 10474 | Third party consultation |
| **Home visit** |  |
| 7883 | Home of Patient |
| 3628 | Out Of Hours Home Visit |
| 7885 | Home Visit |
| 984 | Home Visit - In Surgery Hours |
| 5464 | Home visit note |
| 10627 | Visit-Home |
| 8348 | Normal Home Visit (08:00 - 11:00) |
| 8400 | Nursing Home |
| 9060 | Residential Home |
| 3430 | Night Visit - patient's home |
| 3431 | Night Visit - patient's home |
| 3432 | Night Visit - patient's home |
| 7358 | Daytime Visits patients home |
| 6517 | Seen in own home |
| 3438 | Night Visit patients home |
| 9061 | Residential home visit note |
| 8703 | Nursing home visit note |
| 8527 | Out of hours, Non-Practice |
| 3514 | Nurse Visit |
| 3440 | Night visit, Local rota |
| **Telephone** |  |
| 3506 | Nurse Practitioner Telephone Advice |
| 3640 | Out Of Hours-Telephone Advice |
| 4293 | Telephone Consultation |
| 6731 | Telephone call to relative/carer |
| 3639 | Out Of Hours Telephone Contact/Advice |
| 4289 | Telephone Appt |
| 10359 | Telephone |
| 2527 | Duty Doctor Telephone |
| 3636 | Out Of Hours Telephone Advice |
| 3638 | Out Of Hours Telephone Consultation |

**Supplementary Table 6.** Table of the CPRD Aurum codes and descriptions of consultation types included in the study, grouped by the consultation type category **(Continued)**

| **CPRD Aurum codes: Consultation Source Id** | **Description** |
| --- | --- |
| 5040 | Duty Telephone Appt |
| 6728 | Telephone call from a patient |
| 6730 | Telephone call to a patient |
| 3637 | Out Of Hours Telephone Call |
| 4298 | Telephone encounter |
| 6727 | Telephone Call |
| 9417 | Telephone call from relative/carer |
| 4283 | Telephone Advice |
| 4302 | Telephone Surgery |
| **Triage** |  |
| 9433 | Telephone Triage by Doctor |
| 4303 | Telephone Triage |
| 3510 | Nurse Surgery Triage |
| 4271 | Telephone (Triage) |
| 4394 | Triage By Phone |
| 10510 | Triage |
| 3511 | Nurse Triage |
| 3512 | Nurse Triage Clinic |
| 3513 | Nurse Triage Consultation |
| 3512 | Nurse Triage Clinic |
| 9433 | Telephone Triage by Doctor |
| 4303 | Telephone Triage |
| 6751 | Telephone triage encounter |

**Supplementary Table 7.** Table of costs per length of time for each healthcare professional

| **Healthcare professional** | **2021 cost per length of consultation** | **Source (PSSRU UCHSC*)** |
| --- | --- | --- |
| Physiotherapist | £65 per hour (Band 7) | Chapter 9 (2021) |
| Physiotherapist Specialist Practitioner | £65 per hour (Band 7) |  |
| Physiotherapist Manager | £65 per hour (Band 7) |  |
| Physiotherapist Consultant | £88 per hour (Band 8) |  |
| GP | £34 per 9.22 min | Chapter 10.3b (2021) |
| Nurse GP Practice | £42 per hour | Chapter 10.2 (2021) |
| Staff Nurse | £44 per hour (Band 5) | Chapter 10.1 (2021) |
| Specialist Nurse Practitioner | £66 per hour (Band 7) |  |
| Nurse Manager | £66 per hour (Band 7) |  |
| Nurse Consultant | £146 per hour (Band 9) |  |

*Note: * PSSRU - Personal Social Services Research*

*UCHSC - Unit’s Unit Costs of Health and Social Care*

**Supplementary Table 8.** Table of the length of consultation for each healthcare professional by consultation type

| **Healthcare professional** | **Consultation type** | **Length of consultation** | **Source (PSSRU UCHSC)** |
| --- | --- | --- | --- |
| Physiotherapists | Surgery | 1 hour | Chapter 9 (2021) |
| GP | Surgery | 9.22 min | Chapter 10.3b (2021)  Chapter 10.8a (2012)  Chapter 10.5 (2021) |
|  | Home visit | 11.4 min |  |
|  | Telephone | 7.1 min |  |
|  | Triage | 4 min |  |
| Nurse | Surgery | 15.5 min | Chapter 10.2 (2021)  Chapter 10.7 (2012)  Chapter 10.5 (2021) |
|  | Home visit | 25 min |  |
|  | Telephone | 6.56 min |  |
|  | Triage | 6.56 min |  |

**Supplementary Table 9.** Example of how the cost of a surgery consultation by a Nurse from a GP practice is calculated

**Nurse GP Practice cost per hour = £43**

**Nurse giving a surgery consultation = 15.5 mins**

**Cost for one Nurse from a GP practice giving a surgery consultation**

**= £43/60mins * 15.5 mins = £11.10**

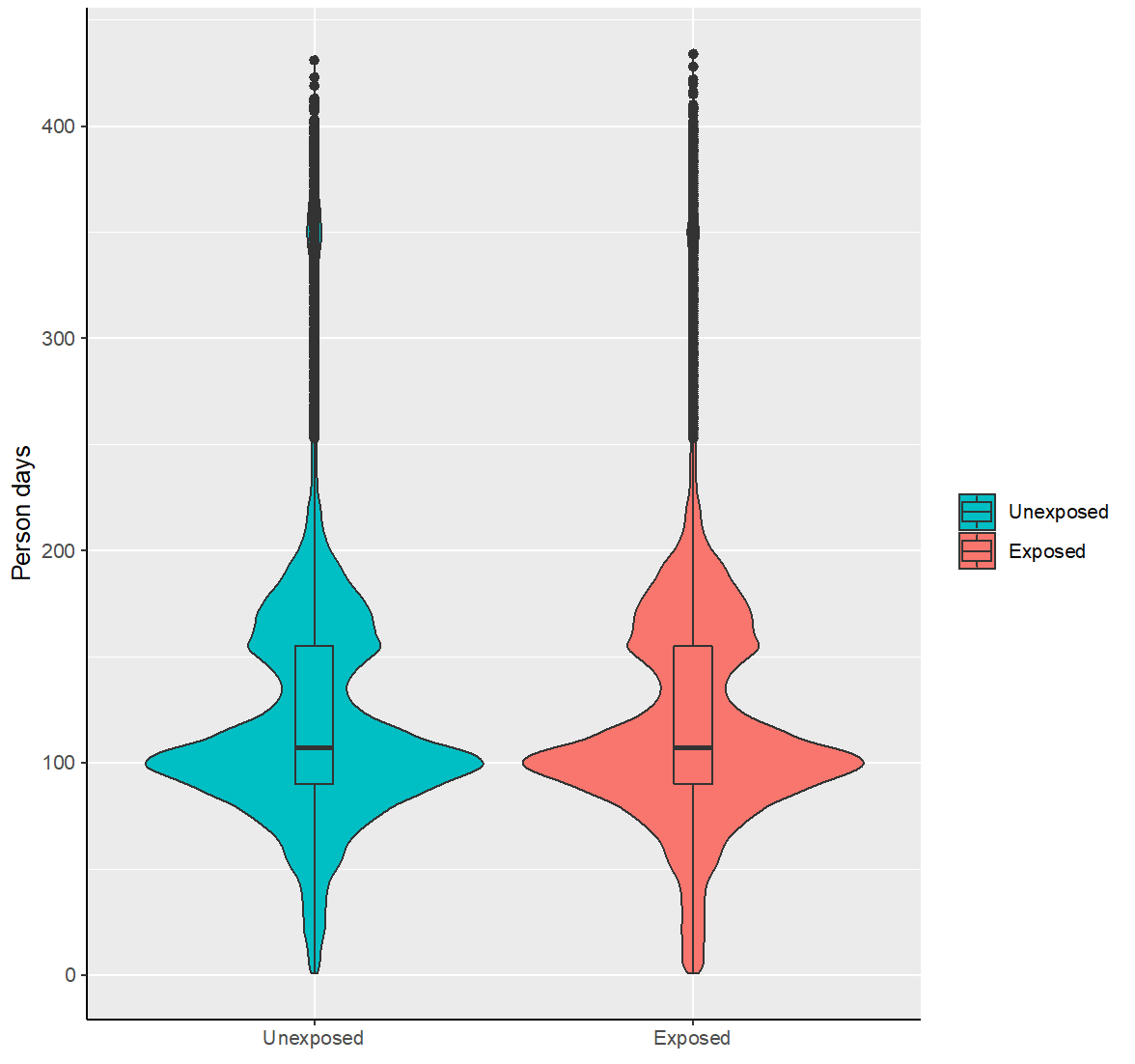

**Supplementary Figure 1.** Violin plot showing the follow up time for the exposed and matched unexposed cohorts.

*Note: The width of each curve represents the frequency of data points in each group. A boxplot has been overlaid to show the mean (thicker line), interquartile ranges (ends of the box) and outliers.*

**Supplementary Figure 2.** Timeline of the dates and time periods of interest throughout the study

31/01/2020 Study start date.

15/04/2020 Start of the time period used to estimate the annual cost of Long COVID.

Index date Date of reported COVID-19 infection for exposed patients, or matched date for unexposed. The earliest index date is 05/02/2022. If the index date is before the 15/04/2020, then patients are prevalent, and if after, they are incident.

12 weeks post index date (Main analysis) Date from which resource use and costs are assumed to be associated with Long COVID, and are included in the analysis.

3 months after 12 weeks post index date (Sensitivity analysis) For those patients who have this data available, the resource use and costs within this time frame are calculated.

15/04/2021 Study end date and end of the time period used to estimate the annual cost of Long COVID.

472,173 eligible exposed patients

8,549,416 unexposed patients

Excluded unexposed patients because:

- Hospitalised within the exclusion timer period.
  - 33,043
- Transferred out of practice before index date.
  - 4,996
- Died before index date.
  - 266
- Transferred out of practice for reasons other than death.
  - 399,152

8,118,797 eligible unexposed patients

472,173 matched unexposed patients

472,173 eligible exposed patients

3,871 exposed patients with a record of diagnosed Long COVID

30,174 exposed patients with reported WHO symptoms of Long COVID after 12 weeks since index date, but without a record of Long COVID diagnosis

472,173 matched exposed patients

Excluded 7,646,624 unexposed patients due to 1:1 propensity score matching

66,303 exposed patients

444 Long COVID diagnosed

10,310 symptoms of Long COVID

64,604 unexposed patients

Excluded 405,870 exposed patients, 407,569 unexposed patients, 3,427 Long COVID diagnosed patients and 19,864 symptoms of Long COVID patients due to them not having 3 months of follow-up after 12 weeks post index date.

**Main analysis**

**Sensitivity analysis**

**Supplementary Figure 3.** Flow chart of the selection of the study population

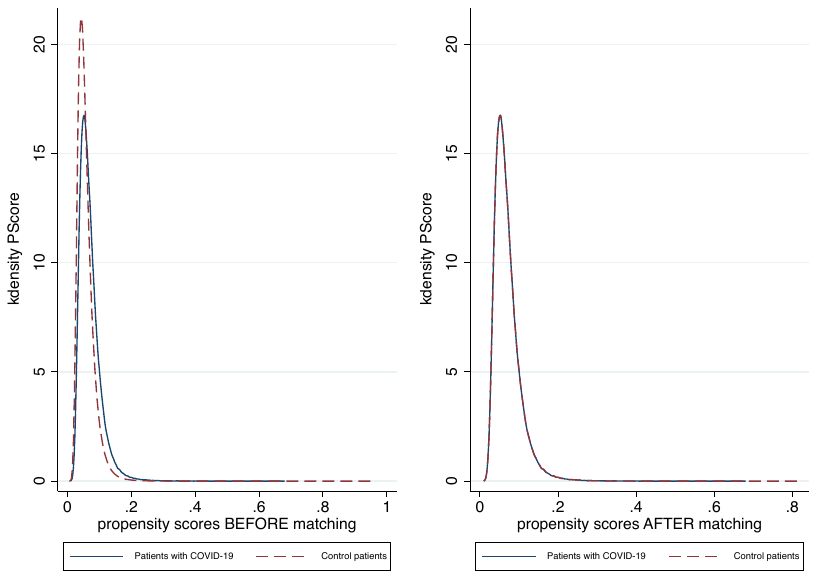

**Supplementary Figure 4**. Kernel density plot before and after propensity score matching to show the comparison in the distribution of the propensity scores between the exposed and unexposed groups

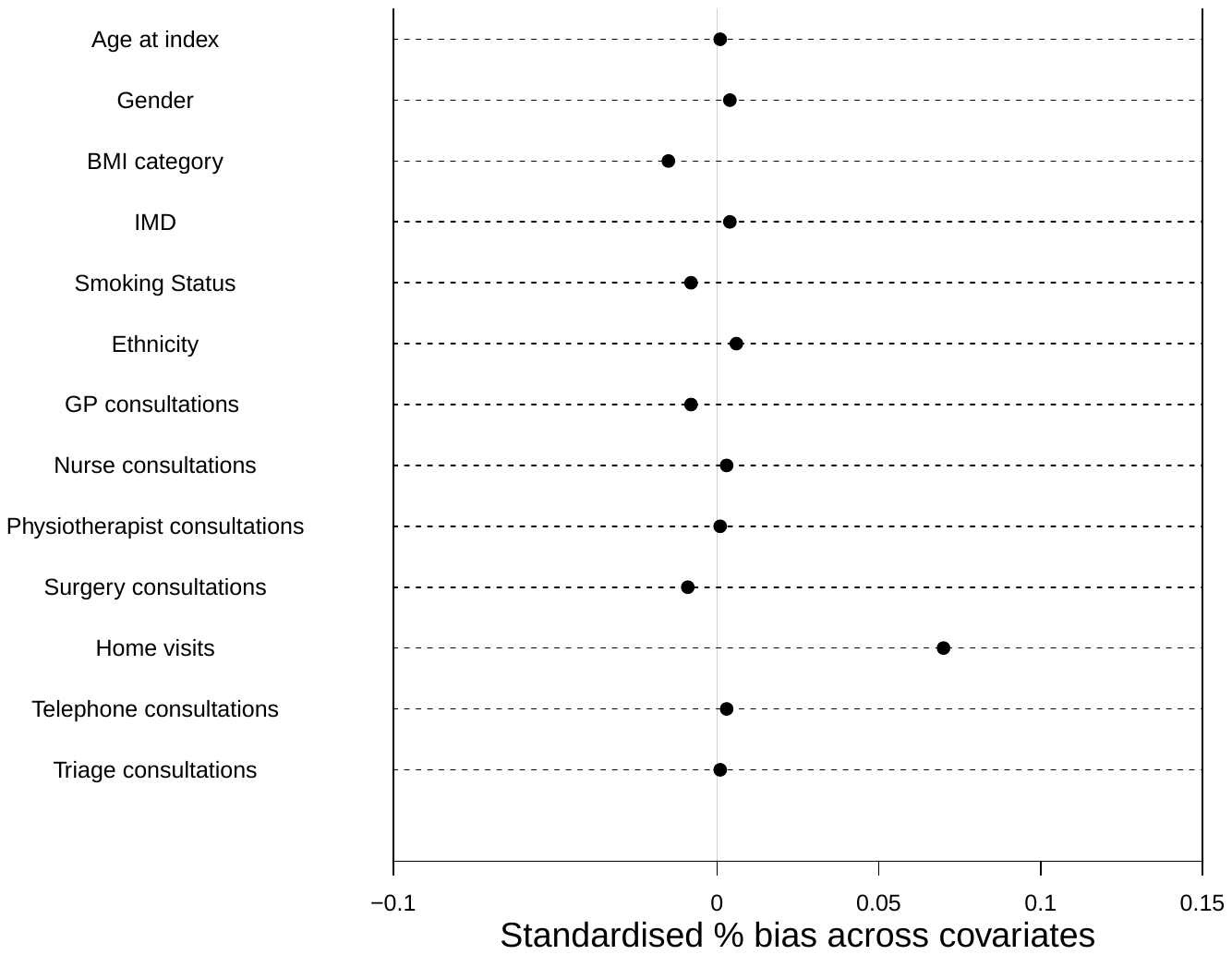

**Supplementary Figure 5.** Standardised differences across covariates in the matched groups

**Supplementary Table 10.** Comorbidities at baseline in matched exposed and unexposed groups

| **Comorbidity** | **Unexposed** | **Exposed** |
| --- | --- | --- |
| Cancer | 20,571 (4.4) | 20,621 (4.4) |
| Arrhythmia | 33,306 (7.1) | 33,575 (7.1) |
| Atrial Fibrillation | 9,185 (1.9) | 9,171 (1.9) |
| Hypertension | 72,412 (15.3) | 71,916 (15.2) |
| Heart Failure | 4,273 (0.9) | 4,176 (0.9) |
| Ischaemic Heart Disease | 13,219 (2.8) | 13,222 (2.8) |
| Myocardial Infarction | 6,599 (1.4) | 6,573 (1.4) |
| Valvular Heart Disease | 5,976 (1.3) | 5,923 (1.3) |
| Cardiomyopathy | 1,236 (0.3) | 1,213 (0.3) |
| Congenital Heart Disease | 2,905 (0.6) | 2,876 (0.6) |
| Peripheral Vascular Disease | 4,346 (0.9) | 4,470 (0.9) |
| Aortic Aneurysm | 960 (0.2) | 919 (0.2) |
| Transient Ischaemic Attack | 4,083 (0.9) | 4,192 (0.9) |
| Ischaemic Stroke | 2,203 (0.5) | 2,297 (0.5) |
| Haemorrhagic Stroke | 1,263 (0.3) | 1,331 (0.3) |
| Stroke Unspecified | 3,549 (0.8) | 3,640 (0.8) |
| Eczema | 92,173 (19.5) | 91,554 (19.4) |
| Psoriasis | 19,491 (4.1) | 19,392 (4.1) |
| Autoimmune Skin Conditions | 6,359 (1.3) | 6,421 (1.4) |
| Acne | 65,310 (13.8) | 64,289 (13.6) |
| Hay Fever | 86,186 (18.3) | 85,419 (18.1) |
| Chronic Sinusitis | 8,306 (1.8) | 8,237 (1.7) |
| Deafness | 5,120 (1.1) | 5,193 (1.1) |
| Blindness | 2,086 (0.4) | 2,159 (0.5) |
| Cataract | 15,743 (3.3) | 16,043 (3.4) |
| Glaucoma | 4,865 (1.0) | 4,955 (1.0) |
| Age-related Macular Degeneration | 3,160 (0.7) | 3,322 (0.7) |
| Diabetic Retinopathy | 12,748 (2.7) | 12,662 (2.7) |
| Inflammatory Eye Disease | 8,073 (1.7) | 8,118 (1.7) |
| Peptic Ulcer | 6,385 (1.4) | 6,426 (1.4) |
| Inflammatory Bowel Disease | 4,767 (1.0) | 4,633 (1.0) |
| Irritable Bowel Syndrome | 33,647 (7.1) | 33,428 (7.1) |
| Hepatitis B | 1,405 (0.3) | 1,362 (0.3) |
| Hepatitis C | 829 (0.2) | 815 (0.2) |
| Alcohol Related Chronic Liver Disease | 852 (0.2) | 860 (0.2) |
| Chronic Liver Disease (All) | 13,489 (2.9) | 13,332 (2.8) |
| Non-Alcoholic Fatty Liver Disease | 6,171 (1.3) | 6,066 (1.3) |
| Diverticular Disease | 13,687 (2.9) | 13,806 (2.9) |

**Supplementary Table 10.** Comorbidities at baseline in matched exposed and unexposed groups **(Continued)**

| **Comorbidity** | **Unexposed** | **Exposed** |
| --- | --- | --- |
| Coeliac Disease | 2,094 (0.4) | 2,066 (0.4) |
| Chronic Pancreatitis | 372 (0.1) | 364 (0.1) |
| Endometriosis | 6,873 (1.5) | 6,916 (1.5) |
| Poly Cystic Ovarian Syndrome | 11,673 (2.5) | 11,667 (2.5) |
| Low Haemoglobin | 25,478 (5.4) | 25,407 (5.4) |
| Venous Thromboembolism | 9,289 (2.0) | 9,181 (1.9) |
| Coagulopathy | 5,463 (1.2) | 5,360 (1.1) |
| Pernicious Anaemia | 1,242 (0.3) | 1,236 (0.3) |
| Depression | 105,548 (22.4) | 104,666 (22.2) |
| Anxiety | 96,512 (20.4) | 96,129 (20.4) |
| Serious Mental Illness | 5,152 (1.1) | 5,167 (1.1) |
| Substance Misuse | 8,275 (1.8) | 8,227 (1.7) |
| Alcohol Misuse | 24,173 (5.1) | 24,162 (5.1) |
| Attention Deficit Hyperactivity Disorder | 2,462 (0.5) | 2,433 (0.5) |
| Eating Disorder | 4,251 (0.9) | 4,187 (0.9) |
| Learning Disability | 4,318 (0.9) | 4,329 (0.9) |
| Alzheimer’s | 3,093 (0.7) | 3,655 (0.8) |
| Vascular Dementia | 1,521 (0.3) | 1,903 (0.4) |
| Dementia Unspecified | 6,768 (1.4) | 7,886 (1.7) |
| Parkinson’s Disease | 893 (0.2) | 945 (0.2) |
| Migraine | 53,130 (1.3) | 52,587 (1.1) |
| Multiple Sclerosis | 1,032 (0.2) | 1,008 (0.2) |
| Epilepsy | 7,495 (1.6) | 7,505 (1.6) |
| Hemiplegia | 710 (0.2) | 703 (0.1) |
| Chronic Fatigue Syndrome | 1,670 (0.4) | 1,667 (0.4) |
| Fibromyalgia | 5,115 (1.1) | 5,027 (1.1) |
| Cluster Headache | 1,758 (0.4) | 1,685 (0.4) |
| Osteoarthritis | 52,342 (11.1) | 52,316 (11.1) |
| Backpain | 6,835 (1.4) | 6,826 (1.4) |
| Fragility Fracture | 45,198 (9.6) | 44,943 (9.5) |
| Falls | 37,672 (8.0) | 38,146 (8.1) |
| Polymyalgia Rheumatica | 2,077 (0.4) | 2,088 (0.4) |
| Rheumatoid Arthritis | 3,991 (0.8) | 4,043 (0.9) |
| Raynaud’s Disease | 5,211 (1.1) | 5,218 (1.1) |
| Sjogren’s Syndrome | 539 (0.1) | 532 (0.1) |
| Systemic Lupus Erythematosus | 600 (0.1) | 592 (0.1) |
| Systemic Sclerosis | 148 (0.0) | 156 (0.0) |
| Ankylosing Spondylitis | 777 (0.2) | 764 (0.2) |

**Supplementary Table 10.** Comorbidities at baseline in matched exposed and unexposed groups **(Continued)**

| **Comorbidity** | **Unexposed** | **Exposed** |
| --- | --- | --- |
| Gout | 13,845 (2.9) | 13,703 (2.9) |
| Chronic Kidney Disease | 13,630 (2.9) | 13,866 (2.9) |
| Asthma | 95,329 (20.2) | 94,787 (20.1) |
| Chronic Obstructive Pulmonary Disease | 10,494 (2.2) | 10,612 (2.2) |
| Obstructive Sleep Apnoea | 7,054 (1.5) | 6,871 (1.5) |
| Other Pulmonary Disease | 2,604 (0.6) | 2,547 (0.5) |
| Hyperthyroidism | 5,226 (1.1) | 5,126 (1.1) |
| Hypothyroidism | 20,720 (4.4) | 20,565 (4.4) |
| Type 1 Diabetes | 2,760 (0.6) | 2,752 (0.6) |
| Type 2 Diabetes | 31,899 (6.8) | 31,673 (6.7) |
| Acquired Immune Deficiency Syndrome | 897 (0.2) | 895 (0.2) |
| Benign Prostatic Hyperplasia | 6,551 (1.4) | 6,574 (1.4) |
| Erectile Dysfunction | 21,407 (4.5) | 20,862 (4.4) |

**Supplementary Table 11.** Incremental cost OLS regression estimates for primary care healthcare cost associated with Long COVID

| **Total healthcare cost** | **Coefficient** | **95% CIs (LL, UL)** | | **p-value** |
| --- | --- | --- | --- | --- |
| Exposure status |  |  |  |  |
| Unexposed (Reference) |  |  |  |  |
| Covid-19 | 2.09 | 1.95 | 2.24 | <0.001 |
| Long Covid diagnosed | 20.50 | 18.50 | 22.50 | <0.001 |
| Symptoms of long Covid | 39.61 | 38.94 | 40.28 | <0.001 |
| Charlson Comorbidity Index | 0.49 | 0.43 | 0.55 | <0.001 |
| GP consultations prior | 2.50 | 2.43 | 2.57 | <0.001 |
| Nurse consultations prior | 1.24 | 1.09 | 1.39 | <0.001 |
| Physio consultations prior | 1.85 | 1.14 | 2.56 | <0.001 |
| Age (at index) | 0.15 | 0.14 | 0.15 | <0.001 |
| Time since index date  Sex |  | 1.81 | 1.85 | <0.001 |
| Male (Reference) |  |  |  |  |
| Female | 1.54 | 1.39 | 1.70 | <0.001 |
| Ethnicity |  |  |  |  |
| White (Reference) |  |  |  |  |
| Black | -0.60 | -0.94 | -0.26 | <0.001 |
| Other | 0.01 | -0.53 | 0.55 | 0.97 |
| Mixed | -0.02 | -0.23 | 0.20 | 0.87 |
| Asian | -0.59 | -1.01 | -0.16 | 0.01 |
| Ethnicity missing | -0.26 | -0.46 | -0.06 | 0.01 |
| IMD |  |  |  |  |
| 1 (Least deprived) (Reference) |  |  |  |  |
| 2 | 0.36 | 0.10 | 0.62 | 0.01 |
| 3 | 0.67 | 0.41 | 0.92 | <0.001 |
| 4 | 0.53 | 0.28 | 0.77 | <0.001 |
| 5 (Most deprived) | 0.82 | 0.56 | 1.07 | <0.001 |
| IMD missing  Smoking status | 0.04 | -0.27 | 0.34 | 0.82 |
| Non-smoker (Reference) |  |  |  |  |
| Ex-smoker | 0.30 | 0.12 | 0.49 | <0.001 |
| Current smoker | 0.14 | -0.05 | 0.33 | 0.14 |
| Smoking status missing | 0.23 | -0.01 | 0.47 | 0.06 |
| BMI category: |  |  |  |  |
| Normal weight (Reference) |  |  |  |  |
| Underweight | 1.48 | 1.08 | 1.88 | <0.001 |
| Overweight | -0.41 | -0.60 | -0.23 | <0.001 |
| Obese | 0.21 | -0.00 | 0.43 | 0.05 |
| BMI missing | 0.65 | 0.41 | 0.89 | <0.001 |
| Constant | -35.27 | -36.07 | -34.47 | <0.001 |

*Notes: R^2^ = 0.297, MSE = 35.642*

**Supplementary Table 12.** Baseline characteristics for the Long COVID subgroups

| **Variables** | **DLC (n = 3,871)** | **SLC (n = 30,172)** |
| --- | --- | --- |
| Age at index (mean (SD)) | 48.41 (12.32) | 45.26 (17.89) |
| Sex |  |  |
| Male | 1,313 (33.9) | 9,204 (30.5) |
| Female | 2,558 (66.1) | 20,968 (69.5) |
| Ethnicity |  |  |
| White | 2,660 (68.7) | 20,294 (67.3) |
| Asian | 464 (12.0) | 3,875 (12.8) |
| Black | 160 (4.1) | 1,039 (3.4) |
| Mixed | 76 (2.0) | 582 (1.9) |
| Other | 55 (1.4) | 437 (1.4) |
| Missing | 456 (11.8) | 3,947 (13.1) |
| IMD |  |  |
| 1 (Least deprived) | 611 (15.8) | 4,060 (13.5) |
| 2 | 642 (16.6) | 4,595 (15.2) |
| 3 | 701 (18.1) | 5,075 (16.8) |
| 4 | 798 (20.6) | 6,109 (20.2) |
| 5 (Most deprived) | 807 (20.8) | 7,514 (24.9) |
| Missing | 312 (8.1) | 2,821 (9.3) |
| Smoking Status |  |  |
| Current smoker | 735 (19.0) | 7,007 (23.2) |
| Ex-Smoker | 1,623 (41.9) | 11,771 (39.0) |
| Never smoked | 1,239 (32.0) | 8,935 (29.6) |
| Missing | 274 (7.1) | 2,461 (8.2) |
| BMI category |  |  |
| Normal weight | 955 (24.7) | 8,927 (29.6) |
| Underweight | 56 (1.4) | 953 (3.2) |
| Obese | 1,477 (38.2) | 9,703 (32.2) |
| Overweight | 1,185 (30.6) | 8,849 (29.3) |
| Missing | 198 (5.1) | 1,742 (5.8) |
| Number of consultations 3 to 12 months prior to index date (mean (SD)) |  |  |
| GP | 3.35 (4.28) | 3.98 (5.00) |
| Nurse | 0.63 (1.43) | 0.89 (2.08) |
| Physiotherapist | 0.02 (0.22) | 0.02 (0.20) |
| Surgery | 1.71 (2.33) | 2.36 (3.09) |
| Home visits | 0.01 (0.13) | 0.16 (1.21) |
| Telephone | 2.25 (3.31) | 2.32 (3.53) |
| Triage | 0.03 (0.30) | 0.04 (0.40) |
| Cancer | 151 (3.9) | 1,681 (5.6) |
| Arrhythmia | 245 (6.3) | 3,168 (10.5) |
| Atrial Fibrillation | 46 (1.2) | 895 (3.0) |
| Hypertension | 735 (19.0) | 5,741 (19.0) |

**Supplementary Table 12.** Baseline characteristics for the Long COVID subgroups **(Continued)**

| **Variables** | **DLC (n = 3,871)** | **SLC (n = 30,172)** |
| --- | --- | --- |
| Heart Failure | 19 (0.5) | 470 (1.6) |
| Ischaemic Heart Disease | 78 (2.0) | 1,419 (4.7) |
| Myocardial Infarction | 31 (0.8) | 651 (2.2) |
| Valvular Heart Disease | 44 (1.1) | 584 (1.9) |
| Cardiomyopathy | 7 (0.2) | 111 (0.4) |
| Congenital Heart Disease | 22 (0.6) | 224 (0.7) |
| Peripheral Vascular Disease | 46 (1.2) | 433 (1.4) |
| Aortic Aneurysm | 6 (0.2) | 83 (0.3) |
| Transient Ischaemic Attack | 48 (1.2) | 447 (1.5) |
| Ischaemic Stroke | 5 (0.1) | 216 (0.7) |
| Haemorrhagic Stroke | 8 (0.2) | 110 (0.4) |
| Stroke Unspecified | 25 (0.6) | 383 (1.3) |
| Eczema | 802 (20.7) | 7,014 (23.2) |
| Psoriasis | 186 (4.8) | 1,497 (5.0) |
| Autoimmune Skin Conditions | 68 (1.8) | 490 (1.6) |
| Acne | 517 (13.4) | 4,900 (16.2) |
| Hay Fever | 847 (21.9) | 6,985 (23.1) |
| Chronic Sinusitis | 141 (3.6) | 873 (2.9) |
| Deafness | 32 (0.8) | 521 (1.7) |
| Blindness | 7 (0.2) | 204 (0.7) |
| Cataract | 75 (1.9) | 1,584 (5.2) |
| Glaucoma | 31 (0.8) | 409 (1.4) |
| Age-related Macular Degeneration | 22 (0.6) | 342 (1.1) |
| Diabetic Retinopathy | 104 (2.7) | 1,014 (3.4) |
| Inflammatory Eye Disease | 96 (2.5) | 645 (2.1) |
| Peptic Ulcer | 59 (1.5) | 669 (2.2) |
| Inflammatory Bowel Disease | 36 (0.9) | 417 (1.4) |
| Irritable Bowel Syndrome | 530 (13.7) | 3,687 (12.2) |
| Hepatitis B | 6 (0.2) | 83 (0.3) |
| Hepatitis C | 6 (0.2) | 72 (0.2) |
| Alcohol Related Chronic Liver Disease | 8 (0.2) | 86 (0.3) |
| Chronic Liver Disease (All) | 203 (5.2) | 1,325 (4.4) |
| Non-Alcoholic Fatty Liver Disease | 112 (2.9) | 632 (2.1) |
| Diverticular Disease | 181 (4.7) | 1,442 (4.8) |
| Coeliac Disease | 19 (0.5) | 209 (0.7) |
| Chronic Pancreatitis | 8 (0.2) | 48 (0.2) |
| Endometriosis | 127 (3.3) | 806 (2.7) |
| Poly Cystic Ovarian Syndrome | 156 (4.0) | 1,169 (3.9) |
| Low Haemoglobin | 302 (7.8) | 2,703 (9.0) |
| Venous Thromboembolism | 119 (3.1) | 932 (3.1) |
| Coagulopathy | 80 (2.1) | 498 (1.7) |
| Pernicious Anaemia | 16 (0.4) | 139 (0.5) |
| Depression | 1,422 (36.7) | 11,366 (37.7) |
| Anxiety | 1,228 (31.7) | 10,598 (35.1) |

**Supplementary Table 12.** Baseline characteristics for the Long COVID subgroups **(Continued)**

| **Variables** | **DLC (n = 3,871)** | **SLC (n = 30,172)** |
| --- | --- | --- |
| Serious Mental Illness | 33 (0.9) | 527 (1.7) |
| Substance Misuse | 69 (1.8) | 791 (2.6) |
| Alcohol Misuse | 205 (5.3) | 1,960 (6.5) |
| Attention Deficit Hyperactivity Disorder | 13 (0.3) | 174 (0.6) |
| Eating Disorder | 55 (1.4) | 514 (1.7) |
| Learning Disability | 9 (0.2) | 305 (1.0) |
| Alzheimer’s | 1 (0.0) | 249 (0.8) |
| Vascular Dementia | 3 (0.1) | 142 (0.5) |
| Dementia Unspecified | 16 (0.4) | 579 (1.9) |
| Parkinson’s Disease | 2 (0.1) | 78 (0.3) |
| Migraine | 767 (19.8) | 5,641 (18.7) |
| Multiple Sclerosis | 12 (0.3) | 98 (0.3) |
| Epilepsy | 55 (1.4) | 592 (2.0) |
| Hemiplegia | 3 (0.1) | 62 (0.2) |
| Chronic Fatigue Syndrome | 59 (1.5) | 214 (0.7) |
| Fibromyalgia | 112 (2.9) | 908 (3.0) |
| Cluster Headache | 26 (0.7) | 170 (0.6) |
| Osteoarthritis | 610 (15.8) | 4,877 (16.2) |
| Backpain | 91 (2.4) | 735 (2.4) |
| Fragility Fracture | 346 (8.9) | 3,284 (10.9) |
| Falls | 377 (9.7) | 3,820 (12.7) |
| Polymyalgia Rheumatica | 13 (0.3) | 236 (0.8) |
| Rheumatoid Arthritis | 30 (0.8) | 384 (1.3) |
| Raynaud’s Disease | 69 (1.8) | 504 (1.7) |
| Sjogren’s Syndrome | 11 (0.3) | 60 (0.2) |
| Systemic Lupus Erythematosus | 8 (0.2) | 65 (0.2) |
| Systemic Sclerosis | 1 (0.0) | 17 (0.1) |
| Ankylosing Spondylitis | 11 (0.3) | 63 (0.2) |
| Gout | 127 (3.3) | 951 (3.2) |
| Chronic Kidney Disease | 78 (2.0) | 1,309 (4.3) |
| Asthma | 1,022 (26.4) | 8,588 (28.5) |
| Chronic Obstructive Pulmonary Disease | 92 (2.4) | 1,749 (5.8) |
| Obstructive Sleep Apnoea | 96 (2.5) | 631 (2.1) |
| Other Pulmonary Disease | 26 (0.7) | 320 (1.1) |
| Hyperthyroidism | 53 (1.4) | 443 (1.5) |
| Hypothyroidism | 271 (7.0) | 1,845 (6.1) |
| Type 1 Diabetes | 13 (0.3) | 208 (0.7) |
| Type 2 Diabetes | 313 (8.1) | 2,587 (8.6) |
| Acquired Immune Deficiency Syndrome | 9 (0.2) | 63 (0.2) |
| Benign Prostatic Hyperplasia | 47 (1.2) | 595 (2.0) |
| Erectile Dysfunction | 199 (5.1) | 1,579 (5.2) |

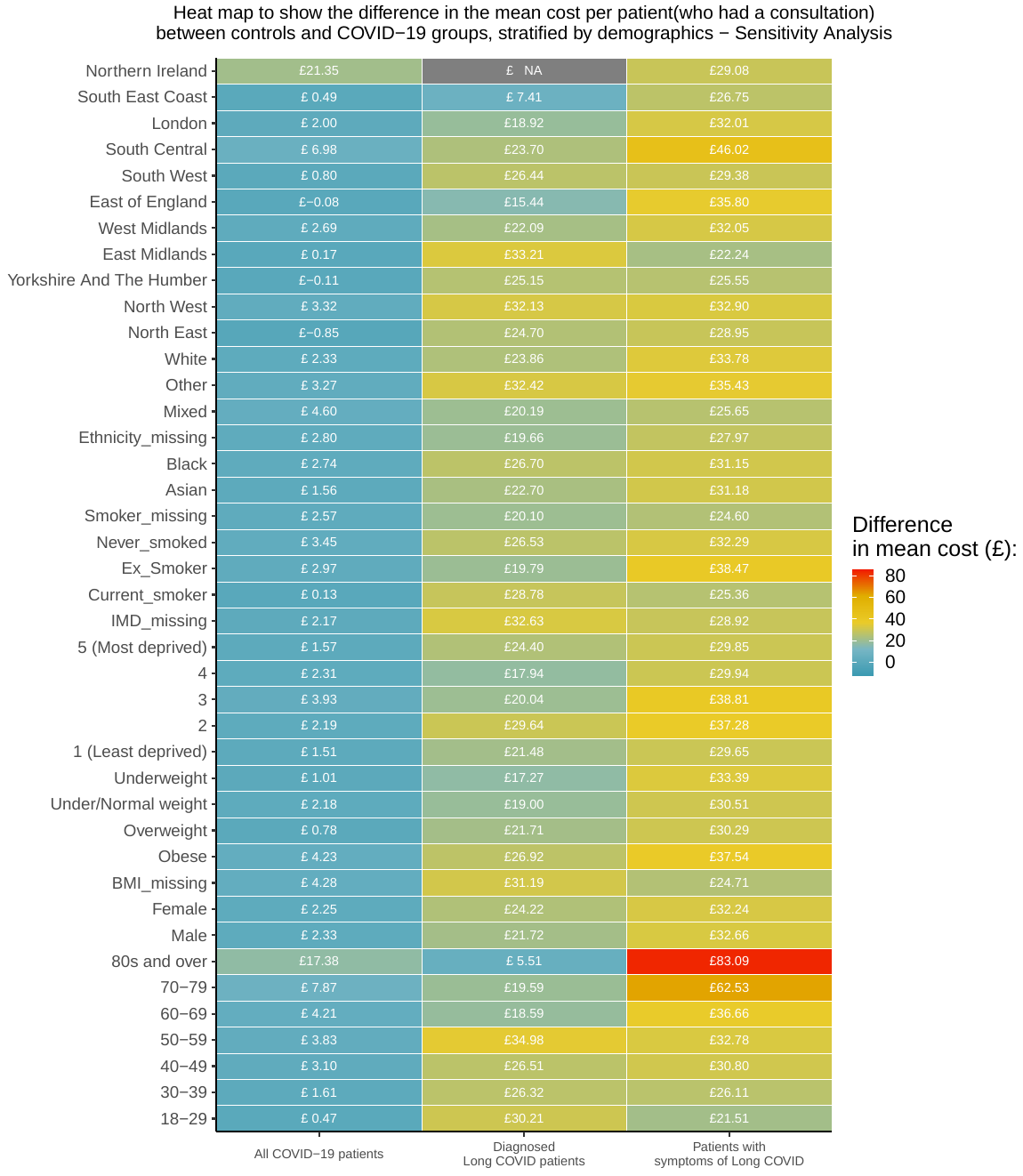

**Supplementary Figure 6.** Heatmap of the difference in mean cost per patient (who had a consultation) between the unexposed patients and COVID-19 groups’ patients, stratified by demographic characteristics

*Note: Yellow and red represent a greater cost difference from the unexposed.*

**Supplementary Table 13.** The mean cost of healthcare professional consultations for COVID-19 patients, stratified by subgroups of patients

|  | **Healthcare professionals – Mean (SD)** | | |  |  |
| --- | --- | --- | --- | --- | --- |
| **Characteristics** | **GP** | **Nurse** | **Physio** | **All** | **p-value *** |
| Age-group |  |  |  |  | < 0.001 |
| 18-29 | 27.87 (3.47) | 11.74 (5.79) | 36.16 (33.15) | 54.23 (51.46) |  |
| 30-39 | 27.96 (3.54) | 11.7 (5.99) | 32.13 (33.74) | 57.13 (60.09) |  |
| 40-49 | 28.03 (3.58) | 11.62 (6.02) | 30.14 (32.91) | 60.42 (64.79) |  |
| 50-59 | 28.09 (3.62) | 11.42 (5.96) | 35.8 (33.48) | 63.46 (70.39) |  |
| 60-69 | 28.22 (3.92) | 11.53 (6.09) | 38.33 (32.58) | 68.04 (80.81) |  |
| 70-79 | 28.89 (4.78) | 12.2 (7.21) | 30.33 (33.04) | 89.08 (120.21) |  |
| 80s and over | 29.71 (5.8) | 13.14 (7.92) | 15.75 (29.8) | 129.74 (150.24) |  |
| Sex |  |  |  |  | < 0.001 |
| Male | 28.34 (4.1) | 11.66 (6.38) | 32.86 (33.29) | 64.44 (78.45) |  |
| Female | 28.24 (4.02) | 11.87 (6.28) | 33.28 (33.5) | 66.98 (79.81) |  |
| Ethnicity |  |  |  |  | < 0.001 |
| White | 28.29 (4.09) | 11.78 (6.33) | 33.41 (33.5) | 67.26 (82.26) |  |
| Asian | 28.15 (3.64) | 12.11 (6.51) | 31.92 (33.29) | 63.56 (65.92) |  |
| Black | 28.15 (3.76) | 11.51 (5.92) | 23.56 (31.84) | 64.04 (73.28) |  |
| Mixed | 28.01 (3.63) | 11.96 (6.06) | 27.58 (32.62) | 62.13 (62.82) |  |
| Other | 28.17 (3.88) | 12.15 (5.73) | 32.5 (32.81) | 63.92 (86.41) |  |
| Missing | 28.41 (4.32) | 11.73 (6.18) | 35.02 (32.94) | 64.21 (79.32) |  |

**Supplementary Table 13.** The mean cost of healthcare professional consultations for COVID-19 patients, stratified by subgroups of patients **(Continued)**

| IMD |  |  |  |  | < 0.001 |
| --- | --- | --- | --- | --- | --- |
| 1 (Least deprived) | 28.25 (4.15) | 11.49 (6.02) | 32.28 (33.24) | 64.01 (80.65) |  |
| 2 | 28.37 (4.19) | 11.55 (6.05) | 30.69 (33.18) | 66.44 (84.72) |  |
| 3 | 28.29 (4.07) | 11.71 (6.14) | 30.93 (32.68) | 68.19 (84.13) |  |
| 4 | 28.22 (3.92) | 11.9 (6.51) | 37.1 (33.03) | 64.99 (76.81) |  |
| 5 (Most deprived) | 28.25 (3.99) | 12.11 (6.44) | 32.51 (34.58) | 67.09 (75.8) |  |
| Missing | 28.28 (3.96) | 11.88 (6.66) | 34.72 (32.8) | 65.39 (72.63) |  |
| Smoking Status |  |  |  |  | < 0.001 |
| Current smoker | 28.2 (3.94) | 11.95 (6.59) | 28.59 (33.17) | 64.48 (75.48) |  |
| Ex-smoker | 28.34 (4.13) | 11.68 (6.29) | 33.74 (33.51) | 70.11 (86.97) |  |
| Never smoked | 28.25 (4) | 11.87 (6.16) | 35.74 (33.05) | 63.47 (73.98) |  |
| Missing | 28.24 (4) | 11.94 (6.14) | 31.72 (33.14) | 61.23 (68.92) |  |
| BMI categories |  |  |  |  | < 0.001 |
| Obese | 28.18 (3.86) | 11.6 (6.14) | 32.78 (33.35) | 70.73 (86.19) |  |
| Overweight | 28.27 (4.02) | 11.78 (6.4) | 32.84 (33.71) | 64.97 (75.88) |  |
| Under/Normal weight | 28.33 (4.14) | 12 (6.38) | 33.92 (32.98) | 63.69 (74.88) |  |
| Underweight | 28.55 (4.63) | 11.87 (6.52) | 28.98 (34.74) | 67.68 (86.23) |  |
| Missing | 28.48 (4.31) | 12.51 (6.4) | 32.85 (33.1) | 59.19 (75.94) |  |
| Region |  |  |  |  | < 0.001 |
| Northeast | 28.68 (4.46) | 10.96 (4.61) | 39.87 (43.01) | 68.68 (73.86) |  |
| Northwest | 28.33 (4.02) | 12.15 (6.56) | 30.49 (32.68) | 70.21 (81.57) |  |
| Yorkshire and the Humber | 27.77 (4.05) | 10.86 (5.21) | 32.67 (35.53) | 69.76 (85.74) |  |
| East Midlands | 27.66 (3.41) | 13.54 (8.36) | 37.49 (33.82) | 67.75 (69.93) |  |
| West Midlands | 28.18 (3.91) | 11.91 (6.57) | 35.24 (32.9) | 66.46 (77.21) |  |
| East of England | 28.11 (4.35) | 12.41 (5.74) | 36.9 (33.45) | 64.34 (75.01) |  |
| Southwest | 28.31 (3.96) | 11.72 (6.89) | 23.05 (31.25) | 66.17 (82.13) |  |
| South Central | 28.34 (4.37) | 10.81 (5.55) | 37.05 (32.85) | 67.41 (88.05) |  |
| London | 28.28 (3.98) | 11.53 (5.99) | 32.33 (33.09) | 60.24 (75.24) |  |
| Southeast Coast | 28.51 (4.06) | 11.53 (5.3) | 39.73 (31.87) | 59.67 (78.74) |  |
| Northern Ireland | 27.49 (5.97) | 11.22 (0.24) | N/A | 77.43 (64.24) |  |

*(*Bootstrapped t-test and ANOVA)*

**Supplementary Table 14.** Estimates of first three months (after index date + 12 weeks) of primary care resource use and costs associated with Long COVID, for patients who had at least three months of follow-up. (Sensitivity analysis)

|  | **Main analysis groups:** | | **COVID-19 patients with:** | | |
| --- | --- | --- | --- | --- | --- |
| **Cost component** | **Unexposed (n = 64,604)** | **Exposed (n = 66,303)** | **DLC  (n = 444)** | | **SLC (n = 10,310)** |
| **Consultations 12 weeks after index date** | | | | | |
| Count | 60,817 | 77,222 | 1,558 | 29,319 | |
| Rate | 0.94 | 1.16 | 3.51 | 2.84 | |
| **Cost (absolute)** | | | | | |
| Total | £1,545,629 | £1,965,131 | £41,059 | £730,980 | |
| Per patient | £23.92 | £29.64 | £92.47 | £70.90 | |
| Per patient who had at least one consultation | £65.34 | £69.52 | £108.62 | £90.24 | |
| **Cost (per person year)** | | | | | |
| Total | £2,507,819 | £3,224,856 | £68,601 | £1,155,146 | |
| Mean | £38.82 | £48.64 | £154.51 | £112.04 | |
| Mean (patients who had at least one consultation) | £105.38 | £113.51 | £180.87 | £142.42 | |

*Notes: DLC – Diagnosed with Long COVID patient group. SLC – Symptoms of Long COVID patient group*

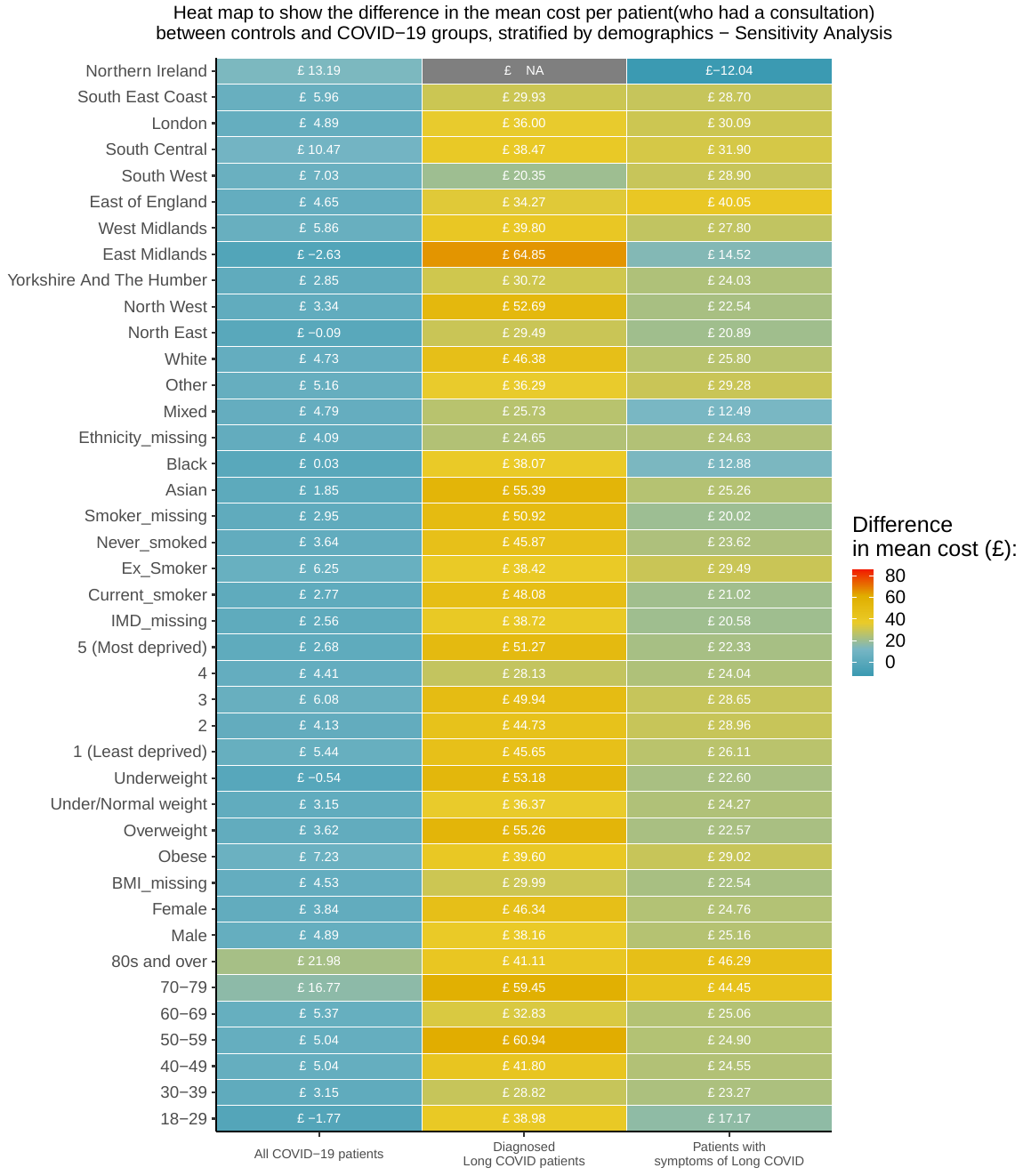

**Supplementary Figure 7.** Heatmap of the difference in mean cost per patient (who had a consultation) between the unexposed patients and COVID-19 groups’ patients, stratified by demographics. – Sensitivity analysis

**Supplementary Table 15.** Table of the number of people estimated by the ONS to have self-reported long COVID between May 2021 and May 2022

| **Region** | **Date** | **Estimated number of people living in private households with self-reported long COVID who first had (or suspected they had) COVID-19 at least 12 weeks previously, UK** |
| --- | --- | --- |
| **East Midlands** | 02/05/2021 | 62 |
|  | 06/06/2021 | 64 |
|  | 04/07/2021 | 63 |
|  | 01/08/2021 | 57 |
|  | 05/09/2021 | 64 |
|  | 02/10/2021 | 63 |
|  | 31/10/2021 | 66 |
|  | 06/12/2021 | 66 |
|  | 02/01/2022 | 76 |
|  | 31/01/2022 | 83 |
|  | 05/03/2022 | 94 |
|  | 03/04/2022 | 93 |
|  | 01/05/2022 | 91 |
| **East of England** | 02/05/2021 | 80 |
|  | 06/06/2021 | 79 |
|  | 04/07/2021 | 75 |
|  | 01/08/2021 | 79 |
|  | 05/09/2021 | 72 |
|  | 02/10/2021 | 67 |
|  | 31/10/2021 | 75 |
|  | 06/12/2021 | 74 |
|  | 02/01/2022 | 82 |
|  | 31/01/2022 | 106 |
|  | 05/03/2022 | 111 |
|  | 03/04/2022 | 126 |
|  | 01/05/2022 | 139 |

**Supplementary Table 15.** Table of the number of people estimated by the ONS to have self-reported long COVID between May 2021 and May 2022 **(Continued)**

| **Region** | **Date** | **Estimated number of people living in private households with self-reported long COVID who first had (or suspected they had) COVID-19 at least 12 weeks previously, UK** |
| --- | --- | --- |
| **London** | 02/05/2021 | 108 |
|  | 06/06/2021 | 106 |
|  | 04/07/2021 | 100 |
|  | 01/08/2021 | 99 |
|  | 05/09/2021 | 108 |
|  | 02/10/2021 | 106 |
|  | 31/10/2021 | 104 |
|  | 06/12/2021 | 103 |
|  | 02/01/2022 | 101 |
|  | 31/01/2022 | 115 |
|  | 05/03/2022 | 122 |
|  | 03/04/2022 | 131 |
|  | 01/05/2022 | 148 |
| **Northeast** | 02/05/2021 | 43 |
|  | 06/06/2021 | 39 |
|  | 04/07/2021 | 39 |
|  | 01/08/2021 | 45 |
|  | 05/09/2021 | 53 |
|  | 02/10/2021 | 45 |
|  | 31/10/2021 | 51 |
|  | 06/12/2021 | 58 |
|  | 02/01/2022 | 59 |
|  | 31/01/2022 | 63 |
|  | 05/03/2022 | 62 |
|  | 03/04/2022 | 70 |
|  | 01/05/2022 | 74 |

**Supplementary Table 15.** Table of the number of people estimated by the ONS to have self-reported long COVID between May 2021 and May 2022 **(Continued)**

| **Region** | **Date** | **Estimated number of people living in private households with self-reported long COVID who first had (or suspected they had) COVID-19 at least 12 weeks previously, UK** |
| --- | --- | --- |
|  | 02/05/2021 | 119 |
|  | 06/06/2021 | 108 |
|  | 04/07/2021 | 117 |
|  | 01/08/2021 | 112 |
|  | 05/09/2021 | 110 |
|  | 02/10/2021 | 118 |
| **Northwest** | 31/10/2021 | 124 |
|  | 06/12/2021 | 112 |
|  | 02/01/2022 | 125 |
|  | 31/01/2022 | 136 |
|  | 05/03/2022 | 159 |
|  | 03/04/2022 | 168 |
|  | 01/05/2022 | 187 |
| **Southeast** | 02/05/2021 | 115 |
|  | 06/06/2021 | 110 |
|  | 04/07/2021 | 111 |
|  | 01/08/2021 | 100 |
|  | 05/09/2021 | 99 |
|  | 02/10/2021 | 103 |
|  | 31/10/2021 | 104 |
|  | 06/12/2021 | 112 |
|  | 02/01/2022 | 117 |
|  | 31/01/2022 | 137 |
|  | 05/03/2022 | 156 |
|  | 03/04/2022 | 173 |
|  | 01/05/2022 | 201 |

**Supplementary Table 15.** Table of the number of people estimated by the ONS to have self-reported long COVID between May 2021 and May 2022 **(Continued)**

| **Region** | **Date** | **Estimated number of people living in private households with self-reported long COVID who first had (or suspected they had) COVID-19 at least 12 weeks previously, UK** |
| --- | --- | --- |
| **Southwest** | 02/05/2021 | 62 |
|  | 06/06/2021 | 59 |
|  | 04/07/2021 | 58 |
|  | 01/08/2021 | 62 |
|  | 05/09/2021 | 67 |
|  | 02/10/2021 | 67 |
|  | 31/10/2021 | 60 |
|  | 06/12/2021 | 63 |
|  | 02/01/2022 | 63 |
|  | 31/01/2022 | 87 |
|  | 05/03/2022 | 101 |
|  | 03/04/2022 | 106 |
|  | 01/05/2022 | 118 |
| **West Midlands** | 02/05/2021 | 70 |
|  | 06/06/2021 | 76 |
|  | 04/07/2021 | 74 |
|  | 01/08/2021 | 75 |
|  | 05/09/2021 | 78 |
|  | 02/10/2021 | 75 |
|  | 31/10/2021 | 85 |
|  | 06/12/2021 | 93 |
|  | 02/01/2022 | 97 |
|  | 31/01/2022 | 96 |
|  | 05/03/2022 | 105 |
|  | 03/04/2022 | 118 |
|  | 01/05/2022 | 128 |

**Supplementary Table 15.** Table of the number of people estimated by the ONS to have self-reported long COVID between May 2021 and May 2022 **(Continued)**

| **Region** | **Date** | **Estimated number of people living in private households with self-reported long COVID who first had (or suspected they had) COVID-19 at least 12 weeks previously, UK** |
| --- | --- | --- |
| **Yorkshire and the Humber** | 02/05/2021 | 81 |
|  | 06/06/2021 | 93 |
|  | 04/07/2021 | 80 |
|  | 01/08/2021 | 78 |
|  | 05/09/2021 | 72 |
|  | 02/10/2021 | 86 |
|  | 31/10/2021 | 79 |
|  | 06/12/2021 | 84 |
|  | 02/01/2022 | 93 |
|  | 31/01/2022 | 103 |
|  | 05/03/2022 | 115 |
|  | 03/04/2022 | 130 |
|  | 01/05/2022 | 147 |
| **Northern Ireland** | 02/05/2021 | 18 |
|  | 06/06/2021 | 16 |
|  | 04/07/2021 | 14 |
|  | 01/08/2021 | 15 |
|  | 05/09/2021 | 15 |
|  | 02/10/2021 | 17 |
|  | 31/10/2021 | 16 |
|  | 06/12/2021 | 20 |
|  | 02/01/2022 | 21 |
|  | 31/01/2022 | 21 |
|  | 05/03/2022 | 23 |
|  | 03/04/2022 | 25 |
|  | 01/05/2022 | 32 |

**Supplementary Table 15.** Table of the number of people estimated by the ONS to have self-reported long COVID between May 2021 and May 2022 **(Continued)**

| **Region** | **Date** | **Estimated number of people living in private households with self-reported long COVID who first had (or suspected they had) COVID-19 at least 12 weeks previously, UK** |
| --- | --- | --- |
| **England** | 02/05/2021 | 741 |
|  | 06/06/2021 | 735 |
|  | 04/07/2021 | 717 |
|  | 01/08/2021 | 706 |
|  | 05/09/2021 | 722 |
|  | 02/10/2021 | 731 |
|  | 31/10/2021 | 747 |
|  | 06/12/2021 | 765 |
|  | 02/01/2022 | 814 |
|  | 31/01/2022 | 925 |
|  | 05/03/2022 | 1024 |
|  | 03/04/2022 | 1115 |
|  | 01/05/2022 | 1233 |

**Supplementary Table 16.** Baseline characteristics for the COVID patients used in the part 2 regression analysis (incurred a positive cost)

| **Variables** | **COVID-19 patients**  **(n = 98,476)** |
| --- | --- |
| Age at index (mean (SD)) | 46.85 (18.10) |
| Gender |  |
| Male | 33,142 (33.7) |
| Female | 65,334 (66.3) |
| BMI category |  |
| Normal weight | 29,383 (29.8) |
| Underweight | 3,052 (3.1) |
| Obese | 29,356 (29.8) |
| Overweight | 30,779 (31.3) |
| Missing | 5,906 (6.0) |
| IMD |  |
| 1 (Least deprived) | 14,839 (15.1) |
| 2 | 15,737 (16.0) |
| 3 | 16,422 (16.7) |
| 4 | 19,702 (20.0) |
| 5 (Most deprived) | 22,617 (23.0) |
| Missing | 9,159 (9.3) |
| Smoking Status |  |
| Current smoker | 20,710 (21.0) |
| Ex-Smoker | 38,929 (39.5) |
| Never smoked | 30,862 (31.3) |
| Missing | 7,975 (8.1) |
| Ethnicity |  |
| White | 66,078 (67.1) |
| Asian | 12,915 (13.1) |
| Black | 3,177 (3.2) |
| Mixed | 1,812 (1.8) |
| Other | 1,239 (1.3) |
| Missing | 13,255 (13.5) |
| Number of consultations 3 to 12 months prior to index date (mean (SD)) |  |
| GP | 3.64 (4.53) |
| Nurse | 0.86 (2.05) |
| Physiotherapist | 0.02 (0.24) |
| Surgery | 2.21 (2.83) |
| Home visits | 0.18 (1.22) |
| Telephone | 2.09 (3.20) |
| Triage | 0.04 (0.37) |

**Supplementary Table 16.** Baseline characteristics for the COVID patients used in the part 2 regression analysis (incurred a positive cost) **(Continued)**

| **Variables** | **COVID-19 patients**  **(n = 98,476)** |
| --- | --- |
| Cancer | 5,941 (6.0) |
| Arrhythmia | 10,437 (10.6) |
| Atrial Fibrillation | 3,100 (3.1) |
| Hypertension | 20,091 (20.4) |
| Heart Failure | 1,457 (1.5) |
| Ischaemic Heart Disease | 4,313 (4.4) |
| Myocardial Infarction | 2,056 (2.1) |
| Valvular Heart Disease | 1,851 (1.9) |
| Cardiomyopathy | 345 (0.4) |
| Congenital Heart Disease | 687 (0.7) |
| Peripheral Vascular Disease | 1,338 (1.4) |
| Aortic Aneurysm | 294 (0.3) |
| Transient Ischaemic Attack | 1,453 (1.5) |
| Ischaemic Stroke | 758 (0.8) |
| Haemorrhagic Stroke | 381 (0.4) |
| Stroke Unspecified | 1,199 (1.2) |
| Eczema | 21,821 (22.2) |
| Psoriasis | 4,823 (4.9) |
| Autoimmune Skin Conditions | 1,446 (1.5) |
| Acne | 14,751 (15.0) |
| Hay Fever | 20,430 (20.7) |
| Chronic Sinusitis | 2,515 (2.6) |
| Deafness | 1,564 (1.6) |
| Blindness | 672 (0.7) |
| Cataract | 5,200 (5.3) |
| Glaucoma | 1,459 (1.5) |
| Age-related Macular Degeneration | 1,121 (1.1) |
| Diabetic Retinopathy | 3,972 (4.0) |
| Inflammatory Eye Disease | 2,150 (2.2) |
| Peptic Ulcer | 1,978 (2.0) |
| Inflammatory Bowel Disease | 1,336 (1.4) |
| Irritable Bowel Syndrome | 10,217 (10.4) |
| Hepatitis B | 258 (0.3) |
| Hepatitis C | 216 (0.2) |
| Alcohol Related Chronic Liver Disease | 251 (0.3) |
| Chronic Liver Disease (All) | 3,961 (4.0) |
| Non-Alcoholic Fatty Liver Disease | 1,835 (1.9) |
| Diverticular Disease | 4,497 (4.6) |
| Coeliac Disease | 588 (0.6) |
| Chronic Pancreatitis | 121 (0.1) |
| Endometriosis | 2,158 (2.2) |

**Supplementary Table 16.** Baseline characteristics for the COVID patients used in the part 2 regression analysis (incurred a positive cost) **(Continued)**

| **Variables** | **COVID-19 patients**  **(n = 98,476)** |
| --- | --- |
| Poly Cystic Ovarian Syndrome | 3,283 (3.3) |
| Low Haemoglobin | 7,982 (8.1) |
| Venous Thromboembolism | 2,916 (3.0) |
| Coagulopathy | 1,511 (1.5) |
| Pernicious Anaemia | 464 (0.5) |
| Depression | 29,482 (29.9) |
| Anxiety | 27,422 (27.8) |
| Serious Mental Illness | 1,682 (1.7) |
| Substance Misuse | 2,073 (2.1) |
| Alcohol Misuse | 5,898 (6.0) |
| Attention Deficit Hyperactivity Disorder | 505 (0.5) |
| Eating Disorder | 1,311 (1.3) |
| Learning Disability | 1,092 (1.1) |
| Alzheimer’s | 1,213 (1.2) |
| Vascular Dementia | 665 (0.7) |
| Dementia Unspecified | 2,613 (2.7) |
| Parkinson’s Disease | 300 (0.3) |
| Migraine | 14,707 (14.9) |
| Multiple Sclerosis | 310 (0.3) |
| Epilepsy | 1,942 (2.0) |
| Hemiplegia | 204 (0.2) |
| Chronic Fatigue Syndrome | 567 (0.6) |
| Fibromyalgia | 1,923 (2.0) |
| Cluster Headache | 468 (0.5) |
| Osteoarthritis | 16,242 (16.5) |
| Backpain | 2140 (2.2) |
| Fragility Fracture | 10,550 (10.7) |
| Falls | 11,472 (11.6) |
| Polymyalgia Rheumatica | 762 (0.8) |
| Rheumatoid Arthritis | 1,320 (1.3) |
| Raynaud’s Disease | 1,462 (1.5) |
| Sjogren’s Syndrome | 196 (0.2) |
| Systemic Lupus Erythematosus | 172 (0.2) |
| Systemic Sclerosis | 47 (0.0) |
| Ankylosing Spondylitis | 205 (0.2) |
| Gout | 3,668 (3.7) |
| Chronic Kidney Disease | 4,612 (4.7) |
| Asthma | 23,792 (24.2) |
| Chronic Obstructive Pulmonary Disease | 3,577 (3.6) |
| Obstructive Sleep Apnoea | 1,992 (2.0) |
| Other Pulmonary Disease | 880 (0.9) |

**Supplementary Table 16.** Baseline characteristics for the COVID patients used in the part 2 regression analysis (incurred a positive cost) **(Continued)**

| **Variables** | **COVID-19 patients**  **(n = 98,476)** |
| --- | --- |
| Hyperthyroidism | 1,508 (1.5) |
| Hypothyroidism | 6,282 (6.4) |
| Type 1 Diabetes | 743 (0.8) |
| Type 2 Diabetes | 9,714 (9.9) |
| Acquired Immune Deficiency Syndrome | 204 (0.2) |
| Benign Prostatic Hyperplasia | 1,968 (2.0) |
| Erectile Dysfunction | 5,371 (5.5) |
